# Social, economic, and environmental disparities in device-measured 24-hour movement behaviours in a nationally representative cohort of older English adults

**DOI:** 10.64898/2026.03.25.26349270

**Authors:** Laura Brocklebank, Andrew Steptoe, Mikaela Bloomberg, Aiden Doherty

## Abstract

**Background:** Insufficient physical activity, excessive sedentary time, and suboptimal sleep are linked to premature mortality and chronic disease and may contribute to social inequalities in health, but most evidence is self-reported. Device-measured, nationally representative data capturing the full 24-hour movement spectrum remain scarce, particularly among older adults. This study examined social, economic, and environmental disparities in 24-hour movement behaviours in the 2021-23 English Longitudinal Study of Ageing (ELSA) accelerometry sub-study.

**Methods:** A subset of 5,382 ELSA participants (71.9%) was invited to wear an Axivity AX3 wrist accelerometer for eight days, with 4,354 (80.9%) agreeing. Raw data were processed using machine learning to derive step count, sleep duration, moderate-to-vigorous and light physical activity, sedentary time, and time in bed. Cross-sectional associations with sex, age, education, marital status, wealth, and urbanicity were examined using linear regression.

**Findings:** Data from 3,648 participants (mean age 68.5 ± 9.3 years; 44.3% men) were included in wear time analyses (median 6.6 days, IQR 6.0-6.9), with 3,161 (86.7%) having sufficient wear time for movement behaviour analyses. Older, unmarried, or lower education/wealth participants were less active, more sedentary, and slept less. Rural participants were more active than urban participants. Women accumulated fewer steps and less moderate-to-vigorous physical activity and sedentary time, but more light activity and longer sleep than men.

**Interpretation:** Social, economic, and environmental disparities exist across the full 24-hour movement spectrum, highlighting population groups for targeted interventions. Follow-up data will clarify how 24-hour movement behaviours influence healthy ageing and contribute to social inequalities in health.

## 1. Introduction

Physical activity, sedentary behaviour, and sleep are interrelated, together making up the full 24-hour movement behaviour profile. Insufficient physical activity, excessive sedentary time, and suboptimal sleep duration are all associated with premature mortality and a higher risk of many chronic diseases.^1,2^ These 24-hour movement behaviours may also reflect the social determinants of health and contribute to social inequalities in health.^3,4^

Since 2004, the World Health Organisation (WHO) has advocated for the systematic monitoring of movement behaviours in national surveillance systems. However, monitoring has largely focused on physical activity rather than the full 24-hour movement behaviour spectrum. Moreover, the most widely used method to date – questionnaires – have several limitations that can lead to highly variable estimates of guideline adherence. These include recall and social desirability biases, inconsistencies in how behaviours are measured, and cultural differences that hinder cross-country comparisons.

Accelerometers can provide more accurate and comparable estimates of movement behaviours. However, there is currently a lack of device-measured movement behaviour data from nationally representative prospective cohort studies,^5^ particularly among older adults, whose behaviours may differ substantially from those of younger and middle-aged adults.^6^ Of the few studies with device data, the most common device placement was the waist. Waist-worn devices typically need to be removed for sleeping, bathing, and swimming, which reduces overall wear time and prevents the capture of the full 24-hour movement behaviour spectrum, particularly sleep. In addition, these studies have mainly relied on proprietary metrics rather than raw acceleration data, limiting harmonisation and comparability.

Raw acceleration data from wrist-worn accelerometers, worn 24 hours per day for 7-9 days, have been collected in several large-scale international epidemiological cohorts to measure movement behaviours.^7–14^ However, only three of these studies were nationally representative: the 2011-14 US National Health and Nutrition Examination Survey (NHANES; aged ≥6 years),^9^ the FinHealth 2017 study (aged ≥25 years),^10^ and the Observation of Cardiovascular Risks and Health in Luxembourg study (ORISCAV-LUX 2; aged ≥18 years).^11^ None of these nationally representative studies processed the raw acceleration data to derive movement behaviours spanning the full 24-hour spectrum. Moreover, while all three examined disparities by age, sex, and education, only one considered broader social and economic factors such as marital status and wealth,^15^ and none examined urbanicity, leaving gaps in our understanding of how 24-hour movement behaviours reflect the social determinants of health and contribute to social inequalities in health.

Therefore, the aim of this study was to examine social, economic, and environmental disparities in 24-hour movement behaviours in the 2021-23 accelerometry sub-study of the nationally representative English Longitudinal Study of Ageing (ELSA).

## 2. Materials and methods

### 2.1. Study population

ELSA is an ongoing nationally representative prospective cohort study of the English population aged ≥50 years. Data collection began in 2002-03 with follow-up every two years. Details of survey design are available elsewhere.^16^ Wave 10 of ELSA received ethical approval from the South Central-Berkshire Research Ethics Committee on March 22, 2021 (21/SC/0030). All participants gave written informed consent, and all procedures were conducted in accordance with the principles of the Declaration of Helsinki.

Between June 2021 and October 2022 (wave 10), approximately 75% of households (including core members and partners) were randomly selected and invited to wear an activity monitor for eight consecutive days and nights to capture their usual levels of physical activity. Subsequently, to mitigate low response due to COVID-19, all households surveyed in the main study between November 2022 and March 2023 were invited to participate.

### 2.2. Data collection

The Axivity AX3 (Axivity Ltd, Newcastle, UK) is a wrist-worn triaxial accelerometer that has been used to measure movement behaviours in other large-scale epidemiological cohorts, such as the UK Biobank (UKB)^7^ and the China Kadoorie Biobank (CKB).^13^ The accelerometers were set to start recording at 10am two working days after postal dispatch and stop recording ten full days later, including three additional days to allow for postal delays. The accelerometers were set to capture triaxial acceleration data at 100 Hz with a dynamic range of ± 8 *g*.

Participants were asked to start wearing the accelerometer immediately after receiving it in the post and wear it on their dominant wrist 24 hours per day for eight consecutive days. Participants were also asked to record on an activity monitoring postcard the date and time when they started wearing the accelerometer. After eight days, participants were asked to mail both the accelerometer and postcard back to the coordinating centre in a pre-paid envelope. In cases where the accelerometer was not returned, the coordinating centre contacted the participant up to four times (by text message, email, or phone call) to confirm its status. Participants were informed that they could wear the waterproof accelerometer when bathing or swimming but not in extremely high temperature or pressure environments (e.g., in a sauna or when diving). Participants were asked to carry on with their normal activities whilst wearing the accelerometer and did not receive feedback on their activity levels until after it was returned.

Information on participants’ social, economic, and environmental factors was collected through computer-assisted telephone, video, or in-person interviews according to prevailing COVID-19 restrictions. Social factors included sex (male or female), age (50-59, 60-69, 70-79, or ≥80 years), and marital status (married or unmarried); economic factors included education (high, intermediate, or low) and household non-housing wealth (quintiles); and environmental factors included urbanicity (urban or rural).

### 2.3. Data processing

Rather than processing the full ten-day monitoring periods, which could have included time when the accelerometer was in transit, we used data from the activity monitoring postcards to define participant-specific seven-day monitoring periods. The start time was set to 00:00 on the day after the participant reported putting the accelerometer on, or 00:00 on the second day of the ten-day monitoring period if postcard data were unavailable. The end time was seven full days after the start time.

The accelerometer-based outcomes considered in the present analysis were total wear time (days), overall daily step count (steps/day), overnight sleep duration (hours/night), time spent in moderate-to-vigorous physical activity (minutes/day), time spent in light physical activity (hours/day), total sedentary time (hours/day), and time spent in bed (hours/day).

#### 2.3.1. Overall daily step count

Overall daily step count (steps/day) was derived using a hybrid machine learning and peak detection algorithm (https://github.com/OxWearables/stepcount, v3.8.0)^17^ and was reported as the median number of steps per day across the seven-day monitoring period. Participants were excluded if the data could not be parsed, the device could not be calibrated, more than 1% of readings were ‘clipped’ (i.e., fell outside ±2 *g*) before or after calibration, or if average acceleration was implausibly high (>100 m*g*). Participants were also excluded if they did not have sufficient wear time (≥3 days of data with coverage across every one-hour period of the 24-hour cycle). Non-wear time was defined as unbroken episodes of ≥90 minutes in which the standard deviation (SD) of acceleration on each axis was <13 m*g*. To account for potential diurnal bias in wear time, recording interruptions and non-wear periods were imputed using the average value from the corresponding minute across the remaining valid days.

#### 2.3.2. Overnight sleep duration

Overnight sleep duration (hours/night) was derived using a sleep staging algorithm (https://github.com/OxWearables/asleep, v0.4.13).^18^ The *asleep* algorithm classifies each 30-second epoch of acceleration data into one of the three sleep stages: 1) wake; 2) rapid eye movement sleep (REM); and 3) non-rapid eye movement sleep (NREM). Overnight sleep duration was defined as the length of the longest sleep window within each noon-to-noon interval, allowing up to 60 minutes of sleep discontinuity, and was reported as the average across the seven-day monitoring period. Participants were excluded if the data could not be parsed, the device could not be calibrated, more than 1% of readings were ‘clipped’ (i.e., fell outside ±3 *g*) before or after calibration, or if average acceleration was implausibly high (>200 m*g*).Participants were also excluded if they did not have sufficient wear time (≥22 hours/day for ≥3 days, including ≥1 weekend day). Non-wear time was defined as unbroken episodes of ≥90 minutes in which the SD of acceleration on each axis was <13 m*g*.

#### 2.3.3. Time spent in 24-hour movement behaviours

Wear time (days), time spent in moderate-to-vigorous physical activity (minutes/day), time spent in light physical activity (hours/day), total sedentary time (hours/day), and time spent in bed (hours/day) were derived using the Biobank Accelerometer Analysis Tool (https://github.com/OxWearables/biobankAccelerometerAnalysis, v7.1.1).^19^ Participants were excluded if the data could not be parsed, the device could not be calibrated, more than 1% of readings were ‘clipped’ (i.e., fell outside ±8 g) before or after calibration, or if average acceleration was implausibly high (>100 m*g*). Participants were also excluded if they did not have sufficient wear time (≥3 days of data with coverage across every one-hour period of the 24-hour cycle). Non-wear time was defined as unbroken episodes of ≥60 minutes in which the SD of acceleration on each axis was <13 m*g*. To account for potential diurnal bias in wear time, recording interruptions and non-wear periods were imputed using the average values from the corresponding minute across the remaining valid days.

### 2.4. Statistical analysis

We first reported the social, economic, and environmental characteristics of the analytic sample, presenting frequency and percentage for categorical variables, mean and SD for normally distributed continuous variables, and median and interquartile range (IQR) for non-normally distributed continuous variables. We also compared the social, economic, and environmental characteristics of individuals who agreed versus declined to wear an accelerometer, with associations assessed using the chi-squared test.

Next, for each accelerometer-based outcome (total wear time, overall daily step count, overnight sleep duration, time spent in moderate-to-vigorous physical activity, time spent in light physical activity, total sedentary time, and time spent in bed), we reported descriptive statistics and examined variation by temporal characteristics (time of day and weekend day versus weekday). Differences by time of day (categorised as 00:00-05:59, 06:00-11:59, 12:00-17:59, or 18:00-23:59) were assessed using repeated measures Analysis of Variance (ANOVA) for normally distributed outcomes and the Friedman test for non-normally distributed outcomes. Differences between weekend days and weekdays were assessed using the paired *t*-test for normally distributed outcomes or the Wilcoxon signed-rank test for non-normally distributed outcomes.

Cross-sectional associations between accelerometer-based outcomes and social, economic, and environmental characteristics were examined using linear regression models. Models for sex, age, and education were adjusted for sex and age group as appropriate (Model 1); models for marital status were additionally adjusted for education (Model 2); models for wealth were further adjusted for marital status (Model 3); and models for urbanicity were additionally adjusted for wealth quintile (Model 4). Marginal means and 95% confidence intervals (CIs) were reported. Linear trends in accelerometer-based outcomes were assessed only for age groups and wealth quintiles, using separate models with age and wealth included as ordinal variables. Finally, to test the representativeness of our results, we conducted a sensitivity analysis restricted to core wave 10 participants with valid accelerometry data, applying the core sample’s wave 10 cross-sectional weights.

Statistical analyses were performed using RStudio (R version 4.4.1). Results have been reported according to the Strengthening the Reporting of Observational Studies in Epidemiology (STROBE) guidelines (Table A1).^20^

## 3. Results

Between June 2021 and March 2023, 7,482 adults aged ≥50 years consented to participate in wave 10 of ELSA, of whom 5,382 (71.9%) were invited to take part in the accelerometry sub-study. Of those invited, 4,354 (80.9%) agreed. The most commonly reported reasons for refusal were concerns about the monitor being uncomfortable (n = 296; 26.5%), the study being too time-consuming (n = 200; 17.9%), and being unable to wear the monitor at work (n = 118; 10.6%).

Figure A1 shows social, economic, and environmental disparities in the percentage of individuals who agreed to wear an accelerometer. As shown in Table A2, age, marital status, education, and wealth were all associated with agreement. Compared with individuals aged 50-59 years, those aged 60-79 years were more likely to agree, whereas those aged ≥80 years were less likely. Individuals who were married, had intermediate or high education, or were in the top two wealth quintiles were also more likely to agree than their respective counterparts.

**Figure 1.**
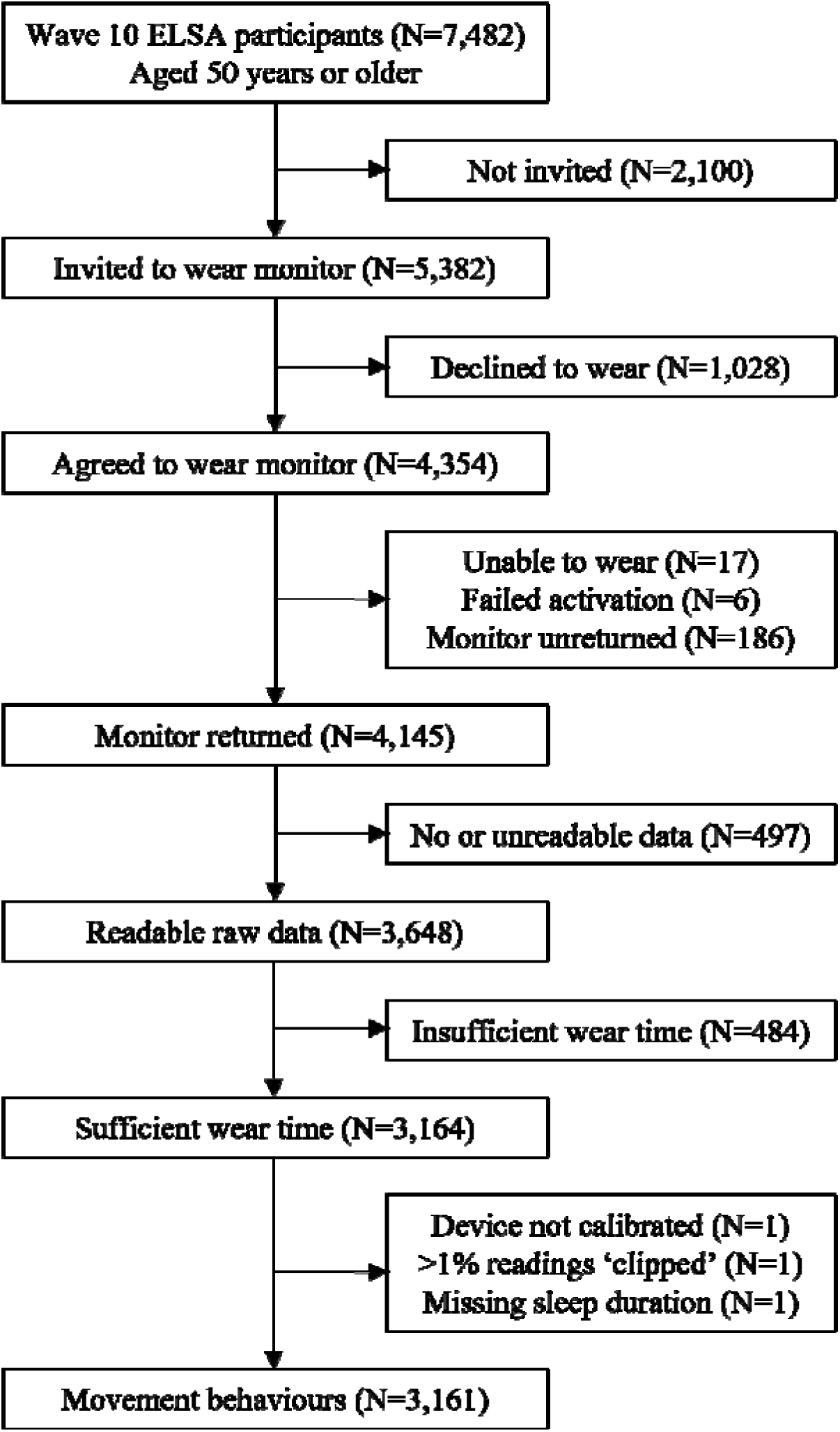
Participant flow chart of the English Longitudinal Study of Ageing (ELSA) accelerometer data collection from 2021-23.

Seventeen participants who agreed to wear an accelerometer were unable to do so due to injury, and a further six devices could not be activated. In total, 4,331 participants were sent an accelerometer by post, of whom 4,145 (95.7%) returned it. Ultimately, 3,648 participants provided readable raw acceleration data and were included in the accelerometry sub-sample.

The social, economic, and environmental characteristics of the accelerometry sub-sample are shown in Table 1. Overall, 2,033 (55.7%) were women and the mean age was 68.5 ± 9.3 years. Most participants lived in an urban area (n = 2,655; 73.0%), were married (n = 2,478; 68.0%), had a high level of education (n = 1,608; 44.7%), and were in the top two wealth quintiles (n = 1,498; 49.4%). The sub-sample was broadly similar to the overall Wave 10 sample.

**Table 1.** Social, economic, and environmental characteristics of the English Longitudinal Study of Ageing (ELSA) accelerometry sub-sample versus the overall Wave 10 sample [N (%)].

| Characteristics | Accelerometry sub-sample | Wave 10 sample |
| --- | --- | --- |
| N (%) | 3648 | 7482 |
| Sex |  |  |
| Men | 1615 (44.3) | 3360 (44.9) |
| Women | 2033 (55.7) | 4122 (55.1) |
| Age (years) |  |  |
| Mean (SD) | 68.5 (9.3) | 68.3 (9.8) |
| 50-59 | 748 (20.5) | 1692 (22.6) |
| 60-69 | 1224 (33.6) | 2502 (33.4) |
| 70-79 | 1206 (33.1) | 2236 (29.9) |
| 80- | 470 (12.9) | 1052 (14.1) |
| Urbanicity <sup>[N=3637/7460]</sup> |  |  |
| Urban | 2655 (73.0) | 5478 (73.4) |
| Rural | 982 (27.0) | 1982 (26.6) |
| Marital status <sup>[N=3646/7477]</sup> |  |  |
| Married | 2478 (68.0) | 4965 (66.4) |
| Not married | 1168 (32.0) | 2512 (33.6) |
| Education <sup>[N=3600/7322]</sup> |  |  |
| High | 1608 (44.7) | 3182 (43.5) |
| Intermediate | 1251 (34.8) | 2487 (34.0) |
| Low | 741 (20.6) | 1653 (22.6) |
| Wealth <sup>[N=3032/6193]</sup> |  |  |
| Quintile 5 (highest) | 747 (24.6) | 1406 (22.7) |
| Quintile 4 (high) | 751 (24.8) | 1470 (23.7) |
| Quintile 3 (intermediate) | 638 (21.0) | 1304 (21.1) |
| Quintile 2 (low) | 500 (16.5) | 1076 (17.4) |
| Quintile 1 (lowest) | 396 (13.1) | 937 (15.1) |

SD, standard deviation.

### 3.1. Wear time

Among the 3,648 participants in the accelerometry sub-sample, the median wear time was 6.6 days (IQR: 6.0-6.9; Table A3). Wear time by temporal characteristics is shown in Table A4. Wear time differed slightly by time of day (*p* < 0.0001), being highest between 12:00-17:59 and lowest between 00:00-05:59 (Figure A2). There was no statistically significant difference in wear time between weekdays and weekend days (*p* = 0.21).

Wear time was generally similar across social, economic, and environmental characteristics (Figure A3 and Table 2), although it was slightly higher among older and wealthier participants. On average, wear time was 3.6 hours/day higher per 10-year older age (*P* for linear trend < 0.0001) and 7.0 hours/day higher in the highest compared with the lowest wealth quintile (*p* = 0.025).

**Table 2.** Social, economic, and environmental disparities in marginal mean (95% CI) wear time and 24-hour movement behaviours.

| Characteristics | Wear time<br>(days) | Step count<br>(steps/day) | Sleep duration<br>(hours/night) | MVPA<br>(minutes/day) | LPA<br>(hours/day) | SB<br>(hours/day) | Time in bed<br>(hours/day) |
| --- | --- | --- | --- | --- | --- | --- | --- |
| N | 3648 | 3161 | 3161 | 3161 | 3161 | 3161 | 3161 |
| Sex |  |  |  |  |  |  |  |
| Men | 5.8<br>(5.7-5.9) | 8155<br>(7921-8389) | 5.9<br>(5.8-6.0) | 35.9<br>(34.1-37.7) | 4.2<br>(4.1-4.3) | 11.8<br>(11.7-12.0) | 7.4<br>(7.3-7.5) |
| Women | 5.7<br>(5.6-5.8) | 7696<br>(7485-7906) | 6.1<br>(6.0-6.1) | 24.5<br>(22.9-26.1) | 4.8<br>(4.7-4.9) | 11.1<br>(11.0-11.2) | 7.7<br>(7.6-7.7) |
| Age (years) |  |  |  |  |  |  |  |
| 50-59 | 5.4<br>(5.3-5.6) | 9809<br>(9463-10154) | 6.3<br>(6.2-6.4) | 39.0<br>(36.4-41.6) | 5.1<br>(4.9-5.2) | 10.7<br>(10.5-10.8) | 7.6<br>(7.5-7.7) |
| 60-69 | 5.7<br>(5.6-5.8) | 9185<br>(8925-9446) | 6.2<br>(6.1-6.3) | 40.3<br>(38.3-42.3) | 4.7<br>(4.6-4.8) | 11.1<br>(11.0-11.3) | 7.5<br>(7.4-7.6) |
| 70-79 | 5.8<br>(5.7-6.0) | 7413<br>(7155-7672) | 6.0<br>(5.9-6.0) | 26.5<br>(24.5-28.4) | 4.4<br>(4.3-4.5) | 11.6<br>(11.5-11.7) | 7.5<br>(7.4-7.6) |
| 80- | 5.9<br>(5.7-6.1) | 5294<br>(4883-5705) | 5.4<br>(5.3-5.6) | 14.9<br>(11.8-18.0) | 3.8<br>(3.7-4.0) | 12.5<br>(12.3-12.7) | 7.5<br>(7.3-7.6) |
| Education <sup>[N=3600/3123]</sup> |  |  |  |  |  |  |  |
| High | 5.7<br>(5.6-5.8) | 8380<br>(8144-8615) | 6.1<br>(6.0-6.1) | 33.9<br>(32.2-35.7) | 4.5<br>(4.4-4.6) | 11.4<br>(11.3-11.5) | 7.5<br>(7.4-7.6) |
| Intermediate | 5.8<br>(5.7-5.9) | 7937<br>(7671-8202) | 6.0<br>(5.9-6.1) | 29.1<br>(27.1-31.1) | 4.6<br>(4.5-4.7) | 11.4<br>(11.3-11.6) | 7.5<br>(7.4-7.6) |
| Low | 5.7<br>(5.5-5.8) | 6993<br>(6657-7329) | 5.8<br>(5.7-5.9) | 24.3<br>(21.7-26.8) | 4.3<br>(4.2-4.5) | 11.7<br>(11.5-11.8) | 7.6<br>(7.5-7.7) |
| Marital status <sup>[N=3598/3122]</sup> |  |  |  |  |  |  |  |
| Married | 5.7<br>(5.7-5.8) | 8030<br>(7826-8235) | 6.0<br>(6.0-6.1) | 29.8<br>(28.2-31.3) | 4.6<br>(4.5-4.7) | 11.3<br>(11.2-11.4) | 7.6<br>(7.5-7.7) |
| Not married | 5.7<br>(5.6-5.8) | 7295<br>(7024-7566) | 5.8<br>(5.7-5.9) | 27.9<br>(25.9-30.00) | 4.3<br>(4.2-4.4) | 11.8<br>(11.6-11.9) | 7.4<br>(7.3-7.5) |
| Wealth <sup>[N=2999/2616]</sup> |  |  |  |  |  |  |  |
| Highest | 5.7<br>(5.5-5.8) | 8627<br>(8266-8989) | 6.0<br>(5.9-6.2) | 35.2<br>(32.5-38.0) | 4.6<br>(4.4-4.8) | 11.2<br>(11.1-11.4) | 7.6<br>(7.5-7.7) |
| High | 5.9<br>(5.7-6.0) | 8092<br>(7755-8429) | 6.1<br>(6.0-6.2) | 32.6<br>(30.1-35.1) | 4.5<br>(4.4-4.7) | 11.3<br>(11.1-11.5) | 7.6<br>(7.5-7.7) |
| Intermediate | 5.8<br>(5.6-5.9) | 7169<br>(6817-7521) | 5.9<br>(5.8-6.0) | 26.0<br>(23.4-28.7) | 4.4<br>(4.3-4.6) | 11.6<br>(11.4-11.8) | 7.5<br>(7.4-7.6) |
| Low | 5.8<br>(5.7-6.0) | 7214<br>(6823-7605) | 5.8<br>(5.7-5.9) | 26.3<br>(23.3-29.2) | 4.3<br>(4.1-4.5) | 11.8<br>(11.6-12.0) | 7.5<br>(7.4-7.6) |
| Lowest | 5.4<br>(5.2-5.6) | 6894<br>(6427-7360) | 5.6<br>(5.4-5.8) | 20.5<br>(17.0-24.0) | 4.4<br>(4.2-4.6) | 12.0<br>(11.8-12.2) | 7.3<br>(7.1-7.4) |
| Urbanicity <sup>[N=2989/2606]</sup> |  |  |  |  |  |  |  |
| Urban | 5.7<br>(5.6-5.8) | 7420<br>(7216-7623) | 5.9<br>(5.8-5.9) | 27.1<br>(25.6-28.6) | 4.4<br>(4.3-4.5) | 11.6<br>(11.5-11.7) | 7.5<br>(7.4-7.6) |
| Rural | 5.8<br>(5.6-5.9) | 8198<br>(7869-8526) | 5.9<br>(5.8-6.1) | 31.6<br>(29.1-34.1) | 4.5<br>(4.4-4.7) | 11.5<br>(11.3-11.6) | 7.5<br>(7.4-7.6) |

Model 1: models for sex, age, and education were adjusted for sex and age group (where appropriate).
Model 2: models for marital status were additionally adjusted for education (Model 1 + education).
Model 3: models for wealth were additionally adjusted for marital status (Model 2 + marital status).
Model 4: models for urbanicity were additionally adjusted for wealth quintile (Model 3 + wealth).
95% CI, 95% confidence interval; MVPA, moderate-to-vigorous physical activity; LPA, light physical activity; SB, sedentary behaviour.

### 3.2. Movement behaviours

Among the accelerometry sub-sample, 3,164 participants (86.7%) had sufficient wear time to assess movement behaviours. A further three participants were excluded because the device could not be calibrated, more than 1% of readings were ‘clipped’ (i.e., fell outside the device’s dynamic range of ±8 *g)* before or after calibration, or one of the outcomes was missing. This resulted in a final analytic sample of 3,161 participants (Figure 1).

#### 3.2.1. Overall daily step count

Among the 3,161 participants with valid movement behaviour data, the median overall daily step count was 7,658 steps/day (IQR: 4,892 to 10,779 steps/day; Table A3). Table A4 shows movement behaviours by temporal characteristics. Median step count was 186 steps/day lower on weekend days than on weekdays (*p* = 0.0027) and differed by time of day (*p* < 0.0001), being highest between 12:00-17:59 and lowest between 00:00-05:59 (Figure A4).

Figure 2 and Table 2 show social, economic, and environmental disparities in overall daily step count. Step count was 459 steps/day lower in women than in men (*p* = 0.0030), 778 steps/day higher in rural compared with urban participants (*p* < 0.0001), and 735 steps/day lower in unmarried compared with married participants (*p* < 0.0001). Step count was, on average, 1,505 steps/day lower per 10-year older age (*P* for linear trend <0.0001). Compared with participants with a high level of education, step count was 443 steps/day lower in those with an intermediate level (*p* = 0.011) and 1,387 steps/day lower in those with a low level (*p* < 0.0001). Finally, step count was, on average, 433 steps/day lower per one-quintile lower wealth (*P* for linear trend <0.0001).

**Figure 2.**
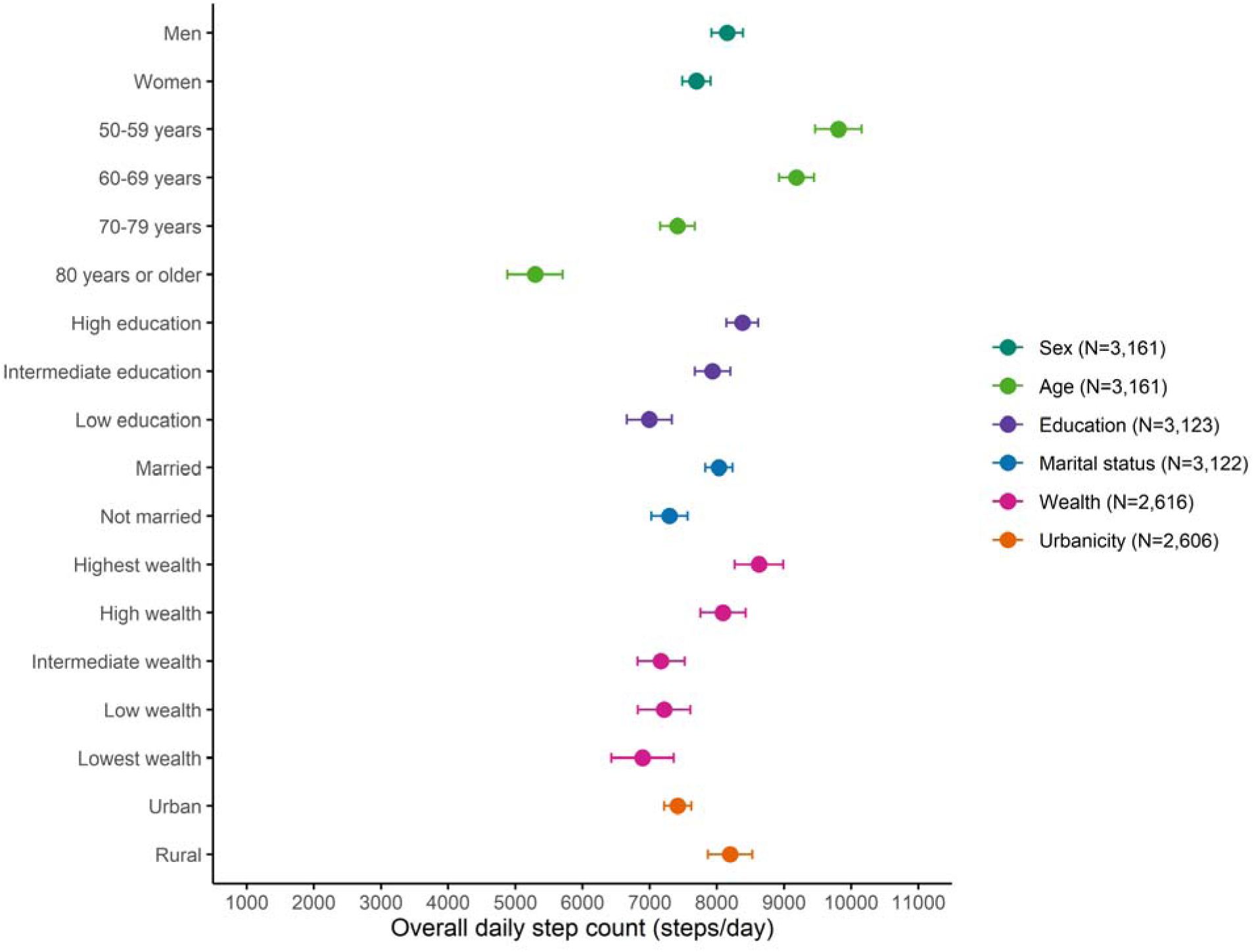
Social, economic, and environmental disparities in overall daily step count (N = 3,161). The markers show marginal means and the horizontal lines show 95% confidence intervals. Model 1: models for sex, age, and education were adjusted for sex and age group (where appropriate). Model 2: models for marital status were additionally adjusted for education (Model 1 + education). Model 3: models for wealth were additionally adjusted for marital status (Model 2 + marital status). Model 4: models for urbanicity were additionally adjusted for wealth quintile (Model 3 + wealth).

#### 3.2.2. Overnight sleep duration

The median overnight sleep duration was 6.3 hours/night (IQR: 5.3-7.0 hours/night; Table A3). Median sleep duration was 6 minutes/day longer on weekend days than on weekdays (*p* < 0.0001; Table A4) and differed by time of day (*p* < 0.0001), being longest between 00:00-05:59 and shortest between 12:00-17:59 (Figure A5).

Figure 3 and Table 2 show social, economic, and environmental disparities in overnight sleep duration. Sleep duration was 11 minutes/night longer in women than in men (*p* < 0.00042) and 15 minutes/night shorter in unmarried compared with married participants (*p* < 0.0001). Sleep duration was, on average, 18 minutes/night shorter per 10-year older age (*P* for linear trend < 0.0001). Compared with participants with a high level of education, sleep duration was 15 minutes/day shorter in those with a low level (*p* < 0.00018) but not significantly different in those with an intermediate level (*p* = 0.15). Sleep duration was, on average, 6 minutes/night shorter per one-quintile lower wealth (*P* for linear trend < 0.0001). There was no statistically significant difference in sleep duration between rural and urban participants (*p* = 0.25).

**Figure 3.**
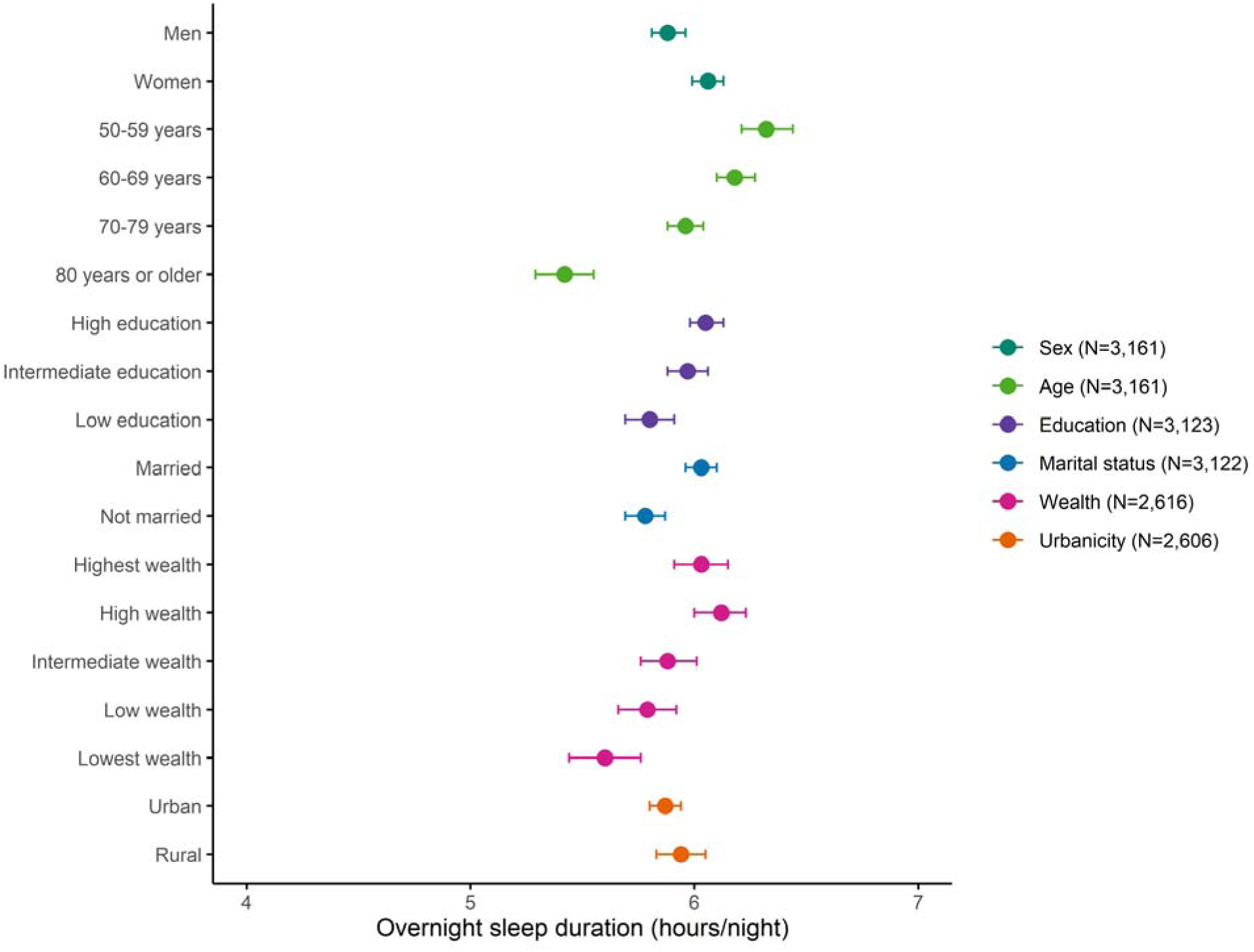
Social, economic, and environmental disparities in overnight sleep duration (N = 3,161). The markers show marginal means and the horizontal lines show 95% confidence intervals. Model 1: models for sex, age, and education were adjusted for sex and age group (where appropriate). Model 2: models for marital status were additionally adjusted for education (Model 1 + education) Model 3: models for wealth were additionally adjusted for marital status (Model 2 + marital status). Model 4: models for urbanicity were additionally adjusted for wealth quintile (Model 3 + wealth).

#### 3.2.3. Time spent in moderate-to-vigorous physical activity

The median amount of time spent in moderate-to-vigorous physical activity was 20.6 minutes/day (IQR: 5.8-45.0 minutes/day; Table A3). Moderate-to-vigorous physical activity differed by time of day (*p* < 0.0001), being highest between 12:00-17:59 and lowest between 00:00-05:59 (Table A4, Figure A6a). There was no statistically significant difference in moderate-to-vigorous physical activity between weekdays and weekend days (*p* = 0.066).

Figure A7 and Table 2 show social, economic, and environmental disparities in time spent in moderate-to-vigorous physical activity. Moderate-to-vigorous physical activity was 11 minutes/day lower in women than in men (*p* < 0.0001) and 5 minutes/day higher in rural compared with urban participants (*p* = 0.0011). Moderate-to-vigorous physical activity was, on average, 8 minutes/day lower per 10-year older age (*P* for linear trend < 0.0001). Compared with participants with a high level of education, MVPA was 5 minutes/day lower in those with an intermediate level (*p* < 0.00024) and 10 minutes/day lower in those with a low level (*p* < 0.0001). Moderate-to-vigorous physical activity was, on average, 4 minutes/day lower per one-quintile lower wealth (*P* for linear trend < 0.0001). There was no statistically significant difference in moderate-to-vigorous physical activity between married and unmarried participants (*p* = 0.15).

#### 3.2.4. Time spent in light physical activity

The median amount of time spent in light physical activity was 4.5 hours/day (IQR: 3.3-5.8 hours/day; Table A3). Median light physical activity was 6 minutes/day lower on weekend days than on weekdays (*p* < 0.0001; Table A4) and differed by time of day (*p* < 0.0001), being highest between 12:00-17:59 and lowest between 00:00-05:59 (Figure A6b).

Figure A8 and Table 2 show social, economic, and environmental disparities in time spent in light physical activity. LPA was 39 minutes/day higher in women than in men (*p* < 0.0001) and 14 minutes/day lower in unmarried compared with married participants (*p* = 0.00077). LPA was, on average, 25 minutes/day lower per 10-year older age (*P* for linear trend < 0.0001) and 3 minutes/day lower per one-quintile lower wealth (*P* for linear trend = 0.0096). There were no statistically significant differences in light physical activity by urbanicity (*p* = 0.23) or education (intermediate: *p* = 0.12; low: *p* = 0.058).

#### 3.2.5. Total sedentary time

Mean total sedentary time was 11.3 hours/day (SD: 2.3 hours/day; Table A3). Mean total sedentary time was 6 minutes/day lower on weekend days than on weekdays (*p* < 0.0001; Table A4) and differed by time of day (*p* < 0.0001), being highest between 18:00-23:59 and lowest between 00:00-05:59 (Figure A6c).

Figure A9 and Table 2 show social, economic, and environmental disparities in total sedentary time. Total sedentary time was 44 minutes/day lower in women than in men (*p* < 0.0001) and 26 minutes/day higher in unmarried compared with married participants (*p* < 0.0001). Total sedentary time was, on average, 36 minutes/day higher per 10-year older age (*P* for linear trend < 0.0001). Compared with participants with a high level of education, total sedentary time was 14 minutes/day higher in those with a low level (*p* = 0.028) but not significantly different in those with an intermediate level (*p* = 0.98). Total sedentary time was, on average, 11 minutes/day higher per one-quintile lower wealth (*P* for linear trend < 0.0001). There was no statistically significant difference in total sedentary time between rural and urban participants (*p* = 0.068).

#### 3.2.6. Time spent in bed

The mean amount of time spent in bed was 7.5 hours/day (SD: 1.3 hours/day; Table A3). Mean time spent in bed was 12 minutes/day higher on weekend days than on weekdays (*p* < 0.0001; Table A4) and differed by time of day (*p* < 0.0001), being highest between 00:00-05:59 and lowest between 12:00-17:59 (Figure A6d).

Figure A10 and Table 2 show social, economic, and environmental disparities in time spent in bed. Time spent in bed was 17 minutes/day higher in women than in men (*p* < 0.0001) and 11 minutes/day lower in unmarried compared with married participants (*p* = 0.00091). Time spent in bed was, on average, 4 minutes/day lower per one-quintile lower wealth (*P* for linear trend = 0.0025). There were no statistically significant differences in time spent in bed by age (*P* for linear trend = 0.17), urbanicity (*p* = 0.89), or education (intermediate: *p* = 0.55; low: *p* = 0.17).

### 3.3. Sensitivity analysis

Social, economic, and environmental disparities were largely unchanged when the core wave 10 cross-sectional weights were applied to the restricted sample (Table A5).

## 4. Discussion

In this accelerometry sub-study of the nationally representative ELSA prospective cohort, response (81%) and adherence (87%) rates were high among English adults aged ≥50 years. Older and wealthier participants wore the accelerometer for longer. Compared with men, women accumulated fewer steps and less moderate-to-vigorous physical activity and sedentary time, but more light activity and longer sleep. Participants who were older, unmarried, or had lower education or wealth were less active, more sedentary, and slept less than their respective counterparts. Rural participants were more active than urban participants. Finally, participants slept longer on weekend days than on weekdays, resulting in less time spent in most waking behaviours.

In terms of socioeconomic disparities in waking behaviours, both lower education and lower wealth were associated with lower physical activity and higher sedentary time. This contrasts with findings from other accelerometry cohorts, which have either reported no significant socioeconomic differences in movement behaviours^10,13,17,19^ or observed higher overall physical activity among participants with lower education or wealth, primarily reflecting more physically demanding (i.e., blue-collar) occupations.^11,15^ Age differences between study populations may partly explain these discrepancies. Participants in our study were older and therefore more likely to be retired, reducing the contribution of occupational physical activity and increasing the relative importance of leisure-time physical activity. Leisure-time physical activity tends to be lower among participants with lower education or wealth due to barriers such as limited energy or financial resources.^11,15^ Socioeconomic disparities in movement behaviours may contribute to health inequalities, reinforcing disadvantaged adults as a key target group for interventions to increase physical activity and reduce sedentary time.

Regarding sex differences in physical activity and sedentary time, our findings are consistent with findings from UKB,^7,17,19^ NHANES,^15^ FinHealth,^10^ ORISCAV-LUX 2,^11^ and CKB,^13^ where overall physical activity was higher in women than in men, largely due to greater engagement in light activities such as household chores and caregiving. In contrast, findings from the Brazilian Study on Nutrition and Health showed a different pattern: women accumulated fewer steps and less moderate-to-vigorous physical activity than men, but this was not offset by a higher ratio of light activity to sedentary behaviour.^21^ This discrepancy may reflect the use of a waist-worn accelerometer, which is less effective at capturing activities of daily living, or the younger age of the participants. Alternatively, it could suggest differences in socioeconomic patterns of movement behaviours between low- to middle-income and high-income countries. Given the limited number of accelerometry cohorts in low- to middle-income countries (LMICs), further research is warranted.

In terms of sleep, we found that participants who were male, older, unmarried, or had lower education or wealth spent less time in bed and had shorter overnight sleep duration. To our knowledge, only UKB and CKB have examined socioeconomic disparities in device-measured sleep.^13,18,19^ Consistent with our findings, UKB reported shorter overnight sleep duration in men, but found a positive rather than inverse association with age.^18^ This discrepancy likely reflects the younger age range in UKB (40-79 years) compared with ELSA (≥50 years), meaning most UKB participants had not yet reached the age at which sleep decline is most pronounced. UKB reported no other socioeconomic differences in overnight sleep duration,^18^ and neither UKB^19^ nor CKB^13^ reported any socioeconomic differences in time spent in bed. Although our findings highlight potential key target groups for sleep interventions, further research is needed in other large-scale international accelerometry cohorts.

### 4.1. Strengths and limitations

A key strength of this study is that the analytic sample was drawn from the nationally representative ELSA cohort, enhancing the generalisability of our findings compared with other wrist-worn accelerometry cohorts such as UKB,^7^ the British Whitehall II study,^8^ the Rotterdam study,^12^ CKB,^13^ and the Pelotas birth cohorts.^14^ Furthermore, unlike nationally representative cohorts from the US,^9^ Finland,^10^ and Luxembourg,^11^ our study focused on older adults – a key target group for movement behaviour interventions given their lower levels of physical activity, higher levels of sedentary time, and shorter sleep duration. However, since not all ELSA participants were invited, the accelerometry sub-sample may not be fully nationally representative. Nonetheless, its characteristics were very similar to those of the full wave 10 sample, and applying the core sample’s wave 10 cross-sectional weights produced largely unchanged results.

Another key strength is that wrist-worn raw acceleration data were processed using three well-validated, open-source machine learning algorithms to derive meaningful movement behaviours across the full 24-hour spectrum. While previous studies have examined socioeconomic disparities in device-measured physical activity, relatively few have investigated such differences in device-measured sleep. However, the algorithms used in the current study were developed using data from younger CAPTURE-24 participants.^17–19^ While the observed social, economic, and environmental disparities support the face validity of these algorithms, further validation in older adults against a ground-truth measure, such as a wearable video camera, is needed.

A final key strength of our study is the examination of disparities across a broader set of factors through which 24-hour movement behaviours both reflect the social determinants of health and contribute to social inequalities in health. While previous studies primarily focused on age, sex, and education, we also examined marital status, wealth, and urbanicity. This broader perspective enabled the identification of additional population groups who may benefit from targeted interventions, specifically unmarried, economically disadvantaged, and urban participants.

Due to the COVID-19 pandemic, data collection was conducted via telephone or video call rather than in person, with accelerometers distributed by post. Nevertheless, both response and adherence rates were high. A limitation of this study is that participants wore the accelerometer for a single seven-day period, which may not fully capture habitual movement behaviours across the year. However, a recent UKB analysis reported moderate-to-good agreement between measurements taken 3-4 years apart, ranging from 58% to 75% across different movement behaviours.^22^ Another limitation is that wrist-worn accelerometers are limited in their ability to detect certain postures or efforts requiring higher energy expenditure, such as carrying loads. Nevertheless, the increasing preference for wrist placement in recent studies, largely driven by higher wear time,^23^ enables ELSA accelerometer data to be pooled and/or compared with other large-scale epidemiological cohorts from the UK,^7,8^ the US,^9^ Europe,^10–12^ China,^13^ and Brazil.^14^

### 4.2. Conclusion

The current study highlights older, unmarried, or socioeconomically disadvantaged adults as key target groups for interventions to increase physical activity, reduce sedentary time, and optimise sleep in later life. Adults living in urban areas may also represent an important target population for physical activity interventions. With follow-up data, the nationally representative ELSA cohort will be well positioned to clarify how movement behaviours across the full 24-hour spectrum influence healthy ageing, reflect the social determinants of health, and contribute to social inequalities in health.

## Acknowledgements

We thank the ELSA study participants for providing the data used in these analyses, and the team at NatCen Social Research for their work on data collection. ChatGPT was used to improve the readability of the manuscript. All authors take full responsibility for the content and interpretation of the manuscript. The authors declare that the results of this study are presented clearly, honestly, and without fabrication, falsification, or inappropriate data manipulation.

## Data availability

Data from ELSA are available to researchers upon application. The interview data used in this study can be accessed free of charge via the UK Data Service (https://ukdataservice.ac.uk/) following registration and agreement to the data’s end user licence. The accelerometer data used in this study will be made available through the UK Data Service in due course.

## Authors’ contributions

Concept and design: LB, AS, and AD. Data access and verification: LB and MB. Statistical analysis: LB. Data interpretation: all authors. Manuscript drafting: LB. Critical revision of the manuscript: all authors. All authors had full access to the data and take final responsibility for the decision to submit the manuscript.

