## Supplementary Material for "Social, economic, and environmental disparities in device-measured 24-hour movement behaviours in a nationally representative cohort of older English adults"

**Table of contents**

Page 2	Table A1. STROBE statement checklist.

Page 3	Table A2. Social, economic, and environmental disparities in the percentage of individuals who agreed to wear an accelerometer (N = 5,382).

Page 4	Table A3. Social, economic, and environmental disparities in wear time and 24-hour movement behaviours.

Page 5	Table A4. Temporal disparities in wear time and 24-hour movement behaviours.

Page 6	Table A5. Social, economic, and environmental disparities in marginal mean (95% CI) wear time and 24-hour movement behaviours after applying the core wave 10 cross-sectional weights to the restricted sample^a^.

Page 7	Figure A1. Social, economic, and environmental disparities in the percentage of individuals who agreed to wear an accelerometer (N = 5,382).

Page 8	Figure A2. 24-hour profile of wear time by age group (N = 3,648).

Page 9	Figure A3. Social, economic, and environmental disparities in wear time (N = 3,648).

Page 10	Figure A4. 24-hour profile of step count by age group (N = 3,161).

Page 11	Figure A5. 24-hour profile of sleep duration by age group (N = 3,161).

Page 12–13	Figure A6. 24-hour profile of MVPA (a), LPA (b), total sedentary time (c), and time spent in bed (d) by age group (N = 3,161).

Page 14	Figure A7. Social, economic, and environmental disparities in time spent in moderate-to-vigorous physical activity (N = 3,161).

Page 15	Figure A8. Social, economic, and environmental disparities in time spent in light physical activity (N = 3,161).

Page 16	Figure A9. Social, economic, and environmental disparities in total sedentary time (N = 3,161).

Page 17	Figure A10. Social, economic, and environmental disparities in time spent in bed (N = 3,161).

**Table A1. STROBE statement checklist.**

|  | **Item number** | **Recommendation** | **Page number** |
| --- | --- | --- | --- |
| **Title and abstract** | 1 | (*a*) Indicate the study’s design with a commonly used term in the title or the abstract | 1,2 |
|  |  | (*b*) Provide in the abstract an informative and balanced summary of what was done and what was found | 2 |
| **Introduction** | | | |
| Background/rationale | 2 | Explain the scientific background and rationale for the investigation being reported | 3-4 |
| Objectives | 3 | State specific objectives, including any prespecified hypotheses | 4 |
| **Methods** | | | |
| Study design | 4 | Present key elements of study design early in the paper | 4 |
| Setting | 5 | Describe the setting, locations, and relevant dates, including periods of recruitment, exposure, follow-up, and data collection | 4 |
| Participants | 6 | (*a*) Give the eligibility criteria, and the sources and methods of selection of participants. Describe methods of follow-up | 4 |
|  |  | (*b*) For matched studies, give matching criteria and number of exposed and unexposed | N/A |
| Variables | 7 | Clearly define all outcomes, exposures, predictors, potential confounders, and effect modifiers. Give diagnostic criteria, if applicable | 5 |
| Data sources/measurement | 8* | For each variable of interest, give sources of data and details of methods of assessment (measurement). Describe comparability of assessment methods if there is more than one group | 4-7 |
| Bias | 9 | Describe any efforts to address potential sources of bias | 4-8 |
| Study size | 10 | Explain how the study size was arrived at | 8,9  Fig 1 |
| Quantitative variables | 11 | Explain how quantitative variables were handled in the analyses. If applicable, describe which groupings were chosen and why | 5 |
| Statistical methods | 12 | (*a*) Describe all statistical methods, including those used to control for confounding | 7-8 |
|  |  | (*b*) Describe any methods used to examine subgroups and interactions | N/A |
|  |  | (*c*) Explain how missing data were addressed | 5-7,9 |
|  |  | (*d*) If applicable, explain how loss to follow-up was addressed | N/A |
|  |  | (*e*) Describe any sensitivity analyses | 7-8 |
| **Results** | | |  |
| Participants | 13* | *(a)* Report numbers of individuals at each stage of study – e.g., numbers potentially eligible, examined for eligibility, confirmed eligible, included in the study, completing follow-up, and analysed | 8,9  Fig 1 |
|  |  | *(b)* Give reasons for non-participation at each stage | 8,9 |
|  |  | *(c)* Consider use of a flow diagram | Fig 1 |
| Descriptive data | 14* | *(a)* Give characteristics of study participants (e.g., demographic, clinical, social) and information on exposures and potential confounders | 8  Table 1 |
|  |  | *(b)* Indicate number of participants with missing data for each variable of interest | Table 1 |
|  |  | *(c)* Summarise follow-up time (e.g., average and total amount) | N/A |
| Outcome data | 15* | Report numbers of outcome events or summary measures over time | N/A |
| Main results | 16 | (*a*) Give unadjusted estimates and, if applicable, confounder-adjusted estimates and their precision (e.g., 95% confidence interval). Make clear which confounders were adjusted for and why they were included | 9-12  Table 2 |
|  |  | (*b*) Report category boundaries when continuous variables were categorised | Table 2 |
|  |  | (*c*) If relevant, consider translating estimates of relative risk into absolute risk for a meaningful time period | N/A |
| Other analyses | 17 | Report other analyses done—e.g., analyses of subgroups and interactions, and sensitivity analyses | 12 |
| **Discussion** |  |  |  |
| Key results | 18 | Summarise key results with reference to study objectives | 12 |
| Limitations | 19 | Discuss limitations of the study, taking into account sources of potential bias or imprecision. Discuss both direction and magnitude of any potential bias | 14-15 |
| Interpretation | 20 | Give a cautious overall interpretation of results considering objectives, limitations, multiplicity of analyses, results from similar studies, and other relevant evidence | 12-14 |
| Generalisability | 21 | Discuss the generalisability (external validity) of the study results | 14 |
| **Other information** |  |  |  |
| Funding | 22 | Give the source of funding and the role of the funders for the present study and, if applicable, for the original study on which the present article is based | 16 |

* Information given separately for exposed and unexposed groups.

**Table A2. Social, economic, and environmental disparities in the percentage of individuals who agreed to wear an accelerometer (N = 5,382).**

| **Characteristics** | **Wear time [N (%)]** | ***P*-value** |
| --- | --- | --- |
| Overall | 4354 (80.9) |  |
| Sex |  | 0.39 |
| Men | 1907 (80.4) |  |
| Women | 2447 (81.3) |  |
| Age (years) |  | **<0.0001** |
| 50-59 | 944 (79.7) |  |
| 60-69 | 1472 (83.1) |  |
| 70-79 | 1387 (83.2) |  |
| 80- | 551 (72.7) |  |
| Urbanicity ^[N=5363]^ |  | 0.10 |
| Urban | 3177 (80.4) |  |
| Rural | 1163 (82.4) |  |
| Marital status ^[N=5377]^ |  | **0.0033** |
| Married | 2909 (82.1) |  |
| Not married | 1443 (78.7) |  |
| Education ^[N=5296]^ |  | **<0.0001** |
| High | 1886 (82.3) |  |
| Intermediate | 1509 (82.7) |  |
| Low | 893 (75.7) |  |
| Wealth ^[N=4528]^ |  | **0.00050** |
| Quintile 5 (highest) | 868 (83.7) |  |
| Quintile 4 (high) | 880 (82.2) |  |
| Quintile 3 (intermediate) | 753 (79.7) |  |
| Quintile 2 (low) | 619 (77.3) |  |
| Quintile 1 (lowest) | 519 (76.9) |  |

**Table A3. Social, economic, and environmental disparities in wear time and 24-hour movement behaviours.**

| **Characteristics** | **Wear time**  **(days)** | **Step count**  **(steps/day)** | **Sleep duration**  **(hours/night)** | **MVPA**  **(minutes/day)** | **LPA**  **(hours/day)** | **SB**  **(hours/day)** | **Time in bed**  **(hours/day)** |
| --- | --- | --- | --- | --- | --- | --- | --- |
| N | 3648 | 3161 | 3161 | 3161 | 3161 | 3161 | 3161 |
| Overall | 6.6  (6.0-6.9) | 7658  (4892-10779) | 6.3  (5.3-7.0) | 20.6  (5.8-45.0) | 4.5  (3.3-5.8) | 11.3 ± 2.3 | 7.5 ± 1.3 |
| Sex |  |  |  |  |  |  |  |
| Men | 6.6  (6.1-7.0) | 7761  (4972-11018) | 6.2  (5.2-6.9) | 25.9  (8.6-55.1) | 4.1  (3.0-5.3) | 11.8 ± 2.3 | 7.4 ± 1.4 |
| Women | 6.5  (5.8-6.9) | 7536  (4839-10506) | 6.4  (5.4-7.0) | 17.1  (4.7-38.7) | 4.8  (3.6-6.1) | 11.0 ± 2.2 | 7.7 ± 1.2 |
| Age (years) |  |  |  |  |  |  |  |
| 50-59 | 6.5  (5.4-6.9) | 9217  (6683-12503) | 6.5  (5.8-7.1) | 29.9  (12.0-53.4) | 5.0  (3.8-6.5) | 10.6 ± 2.2 | 7.6 ± 1.2 |
| 60-69 | 6.6  (6.0-6.9) | 8703  (5580-11967) | 6.4  (5.6-7.1) | 28.6  (10.0-57.9) | 4.6  (3.5-5.9) | 11.1 ± 2.2 | 7.5 ± 1.3 |
| 70-79 | 6.6  (6.2-6.9) | 7076  (4664-9691) | 6.2  (5.3-7.0) | 17.3  (4.9-38.4) | 4.4  (3.2-5.6) | 11.6 ± 2.2 | 7.5 ± 1.4 |
| 80- | 6.6  (6.2-6.9) | 4970  (2626- 7340) | 5.6  (4.4-6.6) | 7.1  (1.2-18.3) | 3.7  (2.7-5.0) | 12.4 ± 2.3 | 7.5 ± 1.6 |
| Urbanicity ^[N=3637/3150]^ |  |  |  |  |  |  |  |
| Urban | 6.5  (5.7-6.9) | 7391  (4688-10356) | 6.3  (5.3-7.0) | 19.6  (5.5-42.4) | 4.4  (3.3-5.8) | 11.4 ± 2.3 | 7.5 ± 1.4 |
| Rural | 6.6  (6.2-6.9) | 8395  (5617-11959) | 6.3  (5.5-7.0) | 24.2  (7.4-54.4) | 4.6  (3.5-5.8) | 11.1 ± 2.2 | 7.5 ± 1.2 |
| Marital status ^[N=3646/3160]^ |  |  |  |  |  |  |  |
| Married | 6.6  (6.1-6.9) | 8108  (5315-11076) | 6.4  (5.5-7.1) | 23.1  (7.2-47.0) | 4.5  (3.4-5.8) | 11.2 ± 2.3 | 7.6 ± 1.3 |
| Not married | 6.5  (5.9-6.9) | 6646  (4018-9707) | 6.1  (5.0-6.8) | 15.8  (4.2-40.9) | 4.4  (3.1-5.6) | 11.6 ± 2.4 | 7.5 ± 1.4 |
| Education ^[N=3600/3123]^ |  |  |  |  |  |  |  |
| High | 6.6  (6.1-6.9) | 8260  (5587- 11098) | 6.3  (5.5-7.0) | 27.4  (9.5-51.2) | 4.4  (3.4-5.7) | 11.3 ± 2.1 | 7.5 ± 1.3 |
| Intermediate | 6.6  (6.0-6.9) | 7749  (4990- 11032) | 6.3  (5.4-7.0) | 19.6  (5.8-43.9) | 4.7  (3.5-6.0) | 11.2 ± 2.4 | 7.5 ± 1.4 |
| Low | 6.5  (5.8-6.9) | 5865  (3352- 9230) | 6.1  (5.0-6.9) | 10.9  (2.1-30.0) | 4.3  (2.9-5.6) | 11.7 ± 2.5 | 7.6 ± 1.4 |
| Wealth ^[N=3032/2641]^ |  |  |  |  |  |  |  |
| Highest | 6.6  (6.1-6.9) | 8811  (6039-11625) | 6.4  (5.5-7.1) | 28.2  (10.9-55.3) | 4.6  (3.5-5.9) | 11.1 ± 2.0 | 7.6 ± 1.2 |
| High | 6.6  (6.3-6.9) | 7643  (5263-10640) | 6.4  (5.6-7.1) | 21.4  (6.9-49.8) | 4.5  (3.4-5.7) | 11.2 ± 2.2 | 7.6 ± 1.3 |
| Intermediate | 6.6  (6.1-6.9) | 6556  (3930-9507) | 6.3  (5.1-7.1) | 14.2  (4.1-34.9) | 4.3  (3.1-5.7) | 11.6 ± 2.4 | 7.5 ± 1.4 |
| Low | 6.6  (6.1-6.9) | 6511  (3999-9252) | 6.1  (5.1-6.8) | 14.1  (2.9-33.5) | 4.3  (3.1-5.4) | 11.7 ± 2.4 | 7.5 ± 1.5 |
| Lowest | 6.5  (5.3-6.7) | 6467  (3899-10161) | 6.0  (5.0-6.8) | 13.0  (3.7-31.6) | 4.4  (3.1-6.1) | 11.7 ± 2.7 | 7.3 ± 1.5 |

Mean ± SD is reported where data were normally distributed, otherwise median (IQR).

MVPA, moderate-to-vigorous physical activity; LPA, light physical activity; SB, sedentary behaviour, SD, standard deviation; IQR, interquartile range.

**Table A4. Temporal disparities in wear time and 24-hour movement behaviours.**

| **Characteristics** | **Wear time**  **(hours/day)** | **Step count**  **(steps/day)** | **Sleep duration**  **(hours/night)** | **MVPA**  **(minutes/day)** | **LPA**  **(hours/day)** | **SB**  **(hours/day)** | **Time in bed**  **(hours/day)** |
| --- | --- | --- | --- | --- | --- | --- | --- |
| N | 3648 | 3161 | 3161 | 3161 | 3161 | 3161 | 3161 |
| Overall | 23.8  (23.3-24.0) | 7658  (4892-10779) | 6.3  (5.3-7.0) | 20.6  (5.8-45.0) | 4.5  (3.3-5.8) | 11.3 ± 2.3 | 7.5 ± 1.3 |
| Weekday | 23.9  (23.4-24.0) | 7642  (4852-10882) | 6.3  (5.3-7.0) | 19.3  (5.2-44.4) | 4.4  (3.3-5.8) | 11.3 ± 2.4 | 7.6 ± 1.3 |
| Weekend | 24.0  (23.4-24.0) | 7456  (4642-10891) | 6.4  (5.2-7.3) | 17.5  (0.0-45.7) | 4.3  (3.1-5.7) | 11.2 ± 2.5 | 7.8 ± 1.5 |
| *P*-value | 0.21 | **0.0027** | **<0.0001** | 0.066 | **<0.0001** | **<0.0001** | **<0.0001** |
| Time of day |  |  |  |  |  |  |  |
| 00:00-05:59 | 5.9  (5.6-6.0) | 51  (16-125) | 4.7  (3.9-5.2) | 0.0  (0.0-0.0) | 0.1  (0.0-0.2) | 0.9 ± 0.9 | 5.0 ± 1.0 |
| 06:00-11:59 | 6.0  (6.0-6.0) | 2754  (1617-4128) | 0.9  (0.4-1.5) | 4.1  (0.0-13.6) | 1.5  (1.0-2.0) | 2.3 ± 0.8 | 2.0 ± 1.0 |
| 12:00-17:59 | 6.0  (6.0-6.0) | 3387  (2108-4896) | 0.0  (0.0-0.0) | 9.8  (2.3-23.4) | 1.8  (1.3-2.4) | 3.7 ± 0.9 | 0.1 ± 0.3 |
| 18:00-23:59 | 6.0  (5.9-6.0) | 1206  (742-1906) | 0.5  (0.1-1.0) | 0.0  (0.0-5.2) | 1.0  (0.6-1.4) | 4.3 ± 0.8 | 0.6 ± 0.6 |
| *P*-value | **<0.0001** | **<0.0001** | **<0.0001** | **<0.0001** | **<0.0001** | **<0.0001** | **<0.0001** |

Mean ± SD is presented where data were normally distributed, otherwise median (IQR).

MVPA, moderate-vigorous physical activity; LPA, light physical activity; SB, sedentary behaviour; SD, standard deviation; IQR, interquartile range.

**Table A5. Social, economic, and environmental disparities in marginal mean (95% CI) wear time and 24-hour movement behaviours after applying the core wave 10 cross-sectional weights to the restricted sample^a^.**

| **Characteristics** | **Wear time**  **(days)** | **Step count**  **(steps/day)** | **Sleep duration**  **(hours/night)** | **MVPA**  **(minutes/day)** | **LPA**  **(hours/day)** | **SB**  **(hours/day)** | **Time in bed**  **(hours/day)** |
| --- | --- | --- | --- | --- | --- | --- | --- |
| N | 3044 | 2650 | 2650 | 2650 | 2650 | 2650 | 2650 |
| Sex |  |  |  |  |  |  |  |
| Men | 5.7  (5.6-5.9) | 7980  (7671-8289) | 5.9  (5.7-6.0) | 34.5  (32.0-37.0) | 4.1  (4.0-4.3) | 11.9  (11.7-12.0) | 7.4  (7.3-7.5) |
| Women | 5.6  (5.5-5.8) | 7458  (7212-7704) | 6.0  (5.9-6.1) | 22.8  (21.2-24.4) | 4.8  (4.7-4.9) | 11.1  (11.0-11.3) | 7.7  (7.6-7.7) |
| Age (years) |  |  |  |  |  |  |  |
| 50-59 | 5.4  (5.2-5.7) | 9804  (9318-10290) | 6.3  (6.1-6.5) | 37.5  (33.7-41.3) | 5.3  (5.0-5.5) | 10.5  (10.3-10.8) | 7.6  (7.4-7.8) |
| 60-69 | 5.6  (5.5-5.8) | 9084  (8681-9486) | 6.2  (6.1-6.3) | 38.2  (35.1-41.3) | 4.7  (4.6-4.9) | 11.1  (11.0-11.3) | 7.5  (7.4-7.6) |
| 70-79 | 5.8  (5.7-6.0) | 7159  (6867-7451) | 6.0  (5.9-6.1) | 25.7  (23.6-27.8) | 4.3  (4.2-4.4) | 11.7  (11.5-11.8) | 7.6  (7.5-7.7) |
| 80- | 5.9  (5.7-6.1) | 4830  (4471-5188) | 5.3  (5.1-5.5) | 13.1  (11.0-15.2) | 3.6  (3.4-3.8) | 12.7  (12.4-12.9) | 7.5  (7.3-7.7) |
| Education ^[N=3013/2626]^ |  |  |  |  |  |  |  |
| High | 5.7  (5.5-5.8) | 8149  (7861-8437) | 6.1  (6.0-6.1) | 32.2  (30.0-34.5) | 4.5  (4.4-4.6) | 11.5  (11.3-11.6) | 7.5  (7.4-7.6) |
| Intermediate | 5.8  (5.6-5.9) | 7923  (7545-8301) | 6.0  (5.9-6.1) | 28.8  (25.9-31.7) | 4.6  (4.4-4.8) | 11.4  (11.2-11.6) | 7.5  (7.4-7.7) |
| Low | 5.6  (5.3-5.8) | 6912  (6509-7316) | 5.7  (5.5-5.9) | 23.7  (20.9-26.5) | 4.3  (4.2-4.5) | 11.7  (11.5-11.9) | 7.6  (7.4-7.7) |
| Marital status ^[N=3011/2625]^ |  |  |  |  |  |  |  |
| Married | 5.7  (5.6-5.9) | 7930  (7662-8199) | 6.0  (5.9-6.1) | 29.4  (27.4-31.4) | 4.6  (4.5-4.7) | 11.3  (11.2-11.5) | 7.6  (7.5-7.7) |
| Not married | 5.6  (5.4-5.8) | 7212  (6877-7547) | 5.7  (5.6-5.9) | 26.3  (23.9-28.7) | 4.3  (4.2-4.5) | 11.8  (11.6-12.0) | 7.4  (7.3-7.6) |
| Wealth ^[N=2978/2595]^ |  |  |  |  |  |  |  |
| Highest | 5.6  (5.4-5.8) | 8580  (8119-9040) | 6.1  (5.9-6.2) | 35.5  (32.0-39.0) | 4.6  (4.4-4.8) | 11.2  (11.0-11.4) | 7.6  (7.5-7.7) |
| High | 5.9  (5.7-6.0) | 8065  (7663-8467) | 6.1  (6.0-6.2) | 32.4  (28.9-35.9) | 4.5  (4.3-4.6) | 11.4  (11.2-11.6) | 7.6  (7.5-7.8) |
| Intermediate | 5.8  (5.6-6.0) | 7153  (6750-7557) | 5.9  (5.8-6.1) | 26.0  (22.7-29.3) | 4.4  (4.2-4.6) | 11.7  (11.4-11.9) | 7.5  (7.4-7.6) |
| Low | 5.7  (5.5-6.0) | 6926  (6391-7460) | 5.7  (5.6-5.9) | 24.1  (20.4-27.9) | 4.2  (4.0-4.4) | 11.9  (11.6-12.1) | 7.5  (7.3-7.7) |
| Lowest | 5.2  (4.9-5.6) | 7208  (6605-7810) | 5.7  (5.4-5.9) | 21.7  (17.9-25.5) | 4.5  (4.2-4.9) | 11.7  (11.4-12.1) | 7.4  (7.1-7.6) |
| Urbanicity ^[N=2978/2595]^ |  |  |  |  |  |  |  |
| Urban | 5.6  (5.5-5.7) | 7388  (7155-7621) | 5.9  (5.8-6.0) | 26.6  (25.0-28.2) | 4.4  (4.3-4.6) | 11.6  (11.4-11.7) | 7.5  (7.4-7.6) |
| Rural | 5.8  (5.6-6.0) | 8238  (7799-8678) | 6.0  (5.8-6.1) | 32.3  (28.7-36.0) | 4.5  (4.3-4.7) | 11.5  (11.3-11.7) | 7.5  (7.3-7.6) |

^a^ Core wave 10 participants with valid accelerometry data.

Model 1: models for sex, age, and education were adjusted for sex and age group (where appropriate).

Model 2: models for marital status were additionally adjusted for education (Model 1 + education).

Model 3: models for wealth were additionally adjusted for marital status (Model 2 + marital status).

Model 4: models for urbanicity were additionally adjusted for wealth quintile (Model 3 + wealth).

95% CI, 95% confidence interval; MVPA, moderate-to-vigorous physical activity; LPA, light physical activity; SB, sedentary behaviour.

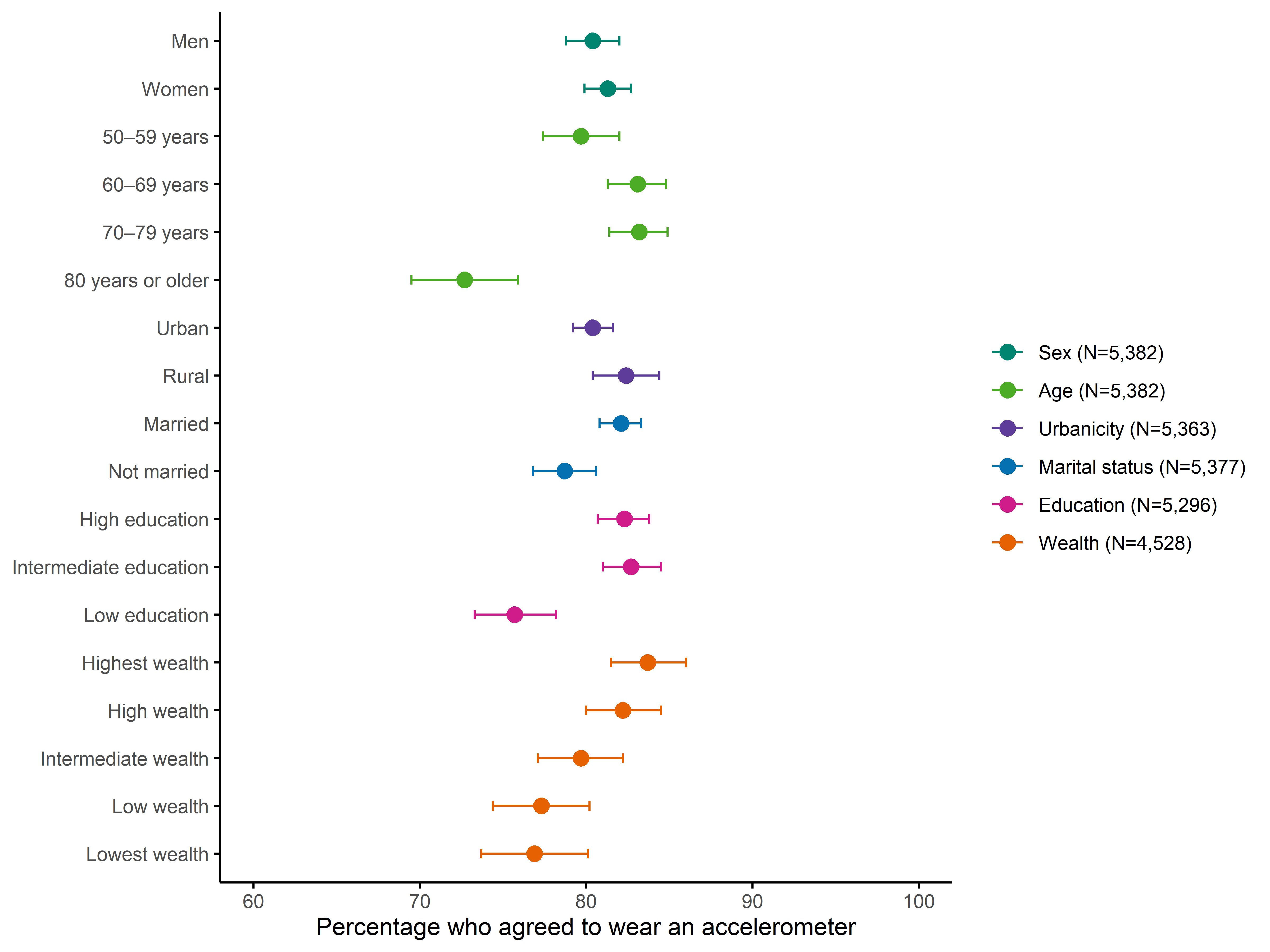
**Figure A1. Social, economic, and environmental disparities in the percentage of individuals who agreed to wear an accelerometer (N = 5,382).**

The horizontal lines show 95% confidence intervals.

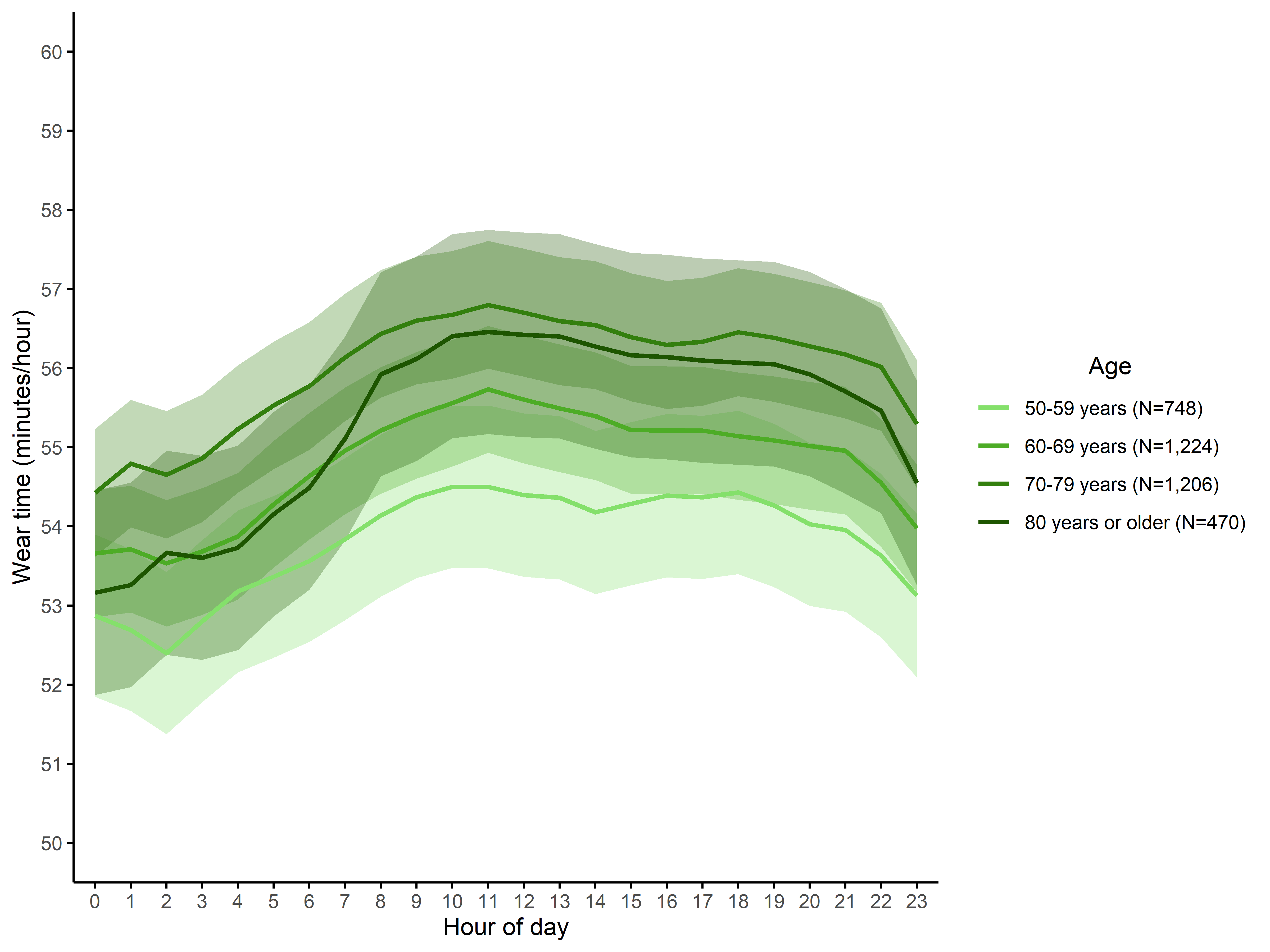
**Figure A2. 24-hour profile of wear time by age group (N = 3,648).**

Means were adjusted for sex and ribbon represents the 95% confidence interval.

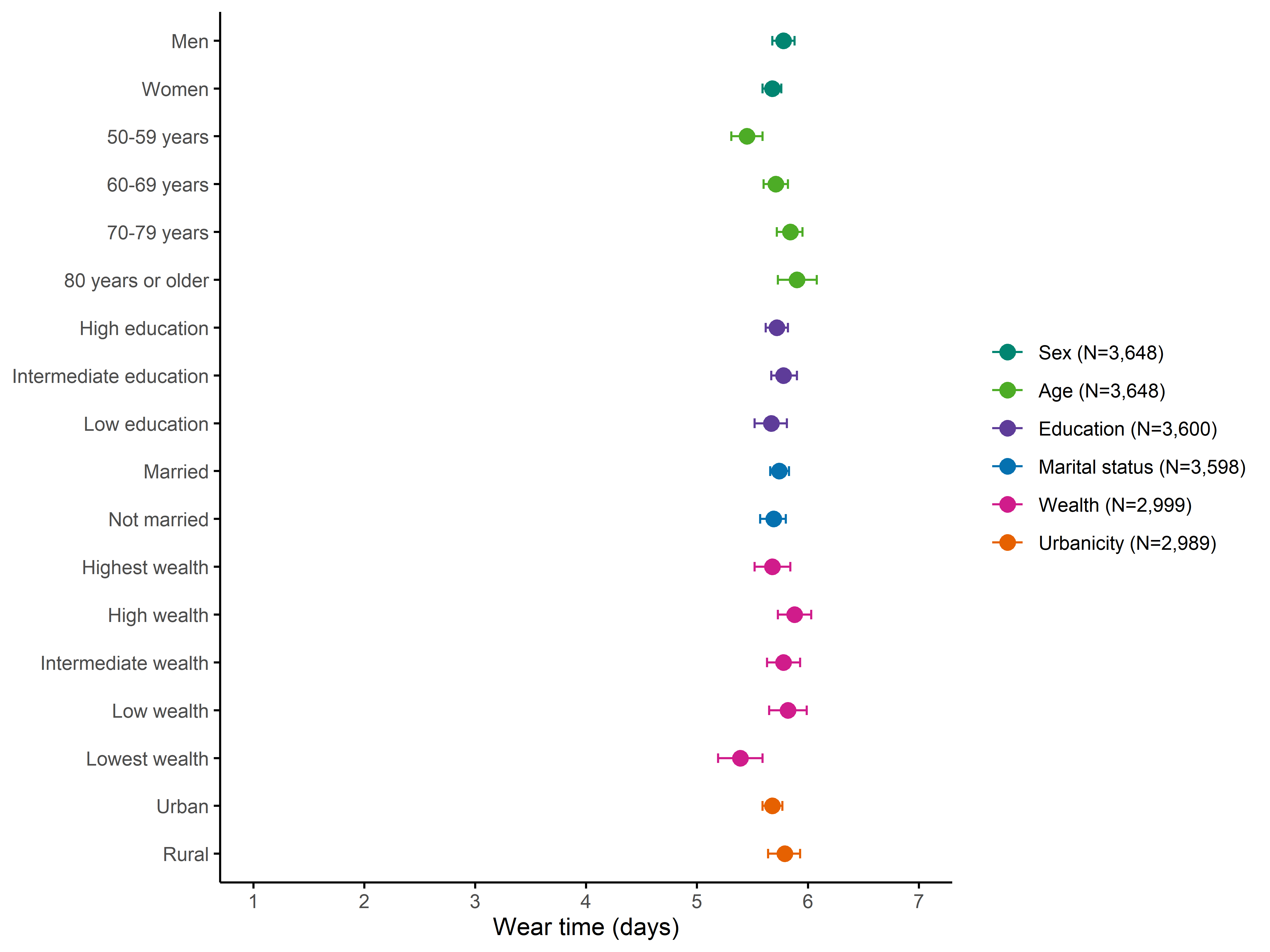
**Figure A3. Social, economic, and environmental disparities in wear time (N = 3,648).**

The markers show marginal means and the horizontal lines show 95% confidence intervals.

Model 1: models for sex, age, and education were adjusted for sex and age group (where appropriate).

Model 2: models for marital status were additionally adjusted for education (Model 1 + education).

Model 3: models for wealth were additionally adjusted for marital status (Model 2 + marital status).

Model 4: models for urbanicity were additionally adjusted for wealth quintile (Model 3 + wealth).

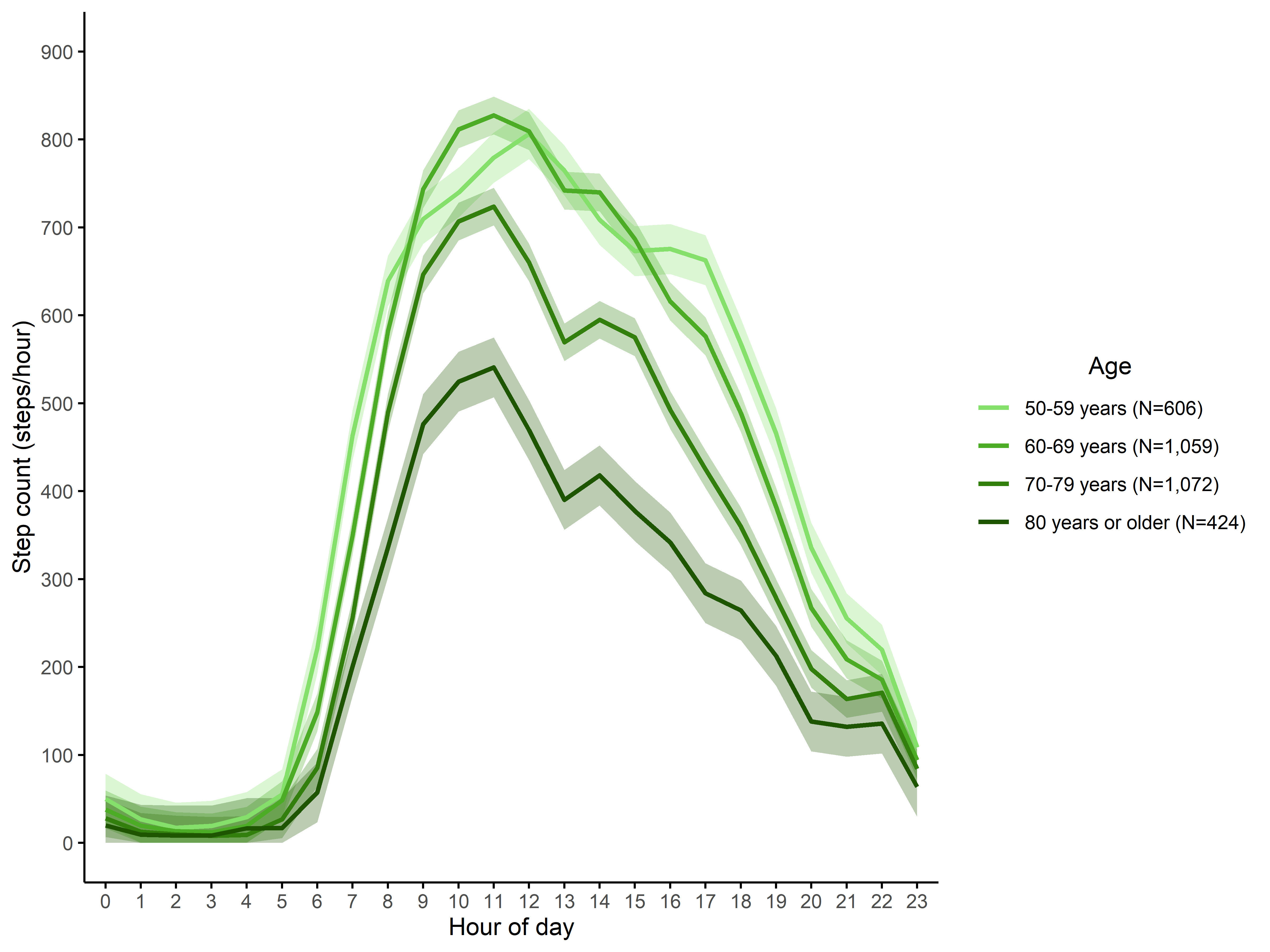
**Figure A4. 24-hour profile of step count by age group (N = 3,161).**

Means were adjusted for sex and ribbon represents the 95% confidence interval.

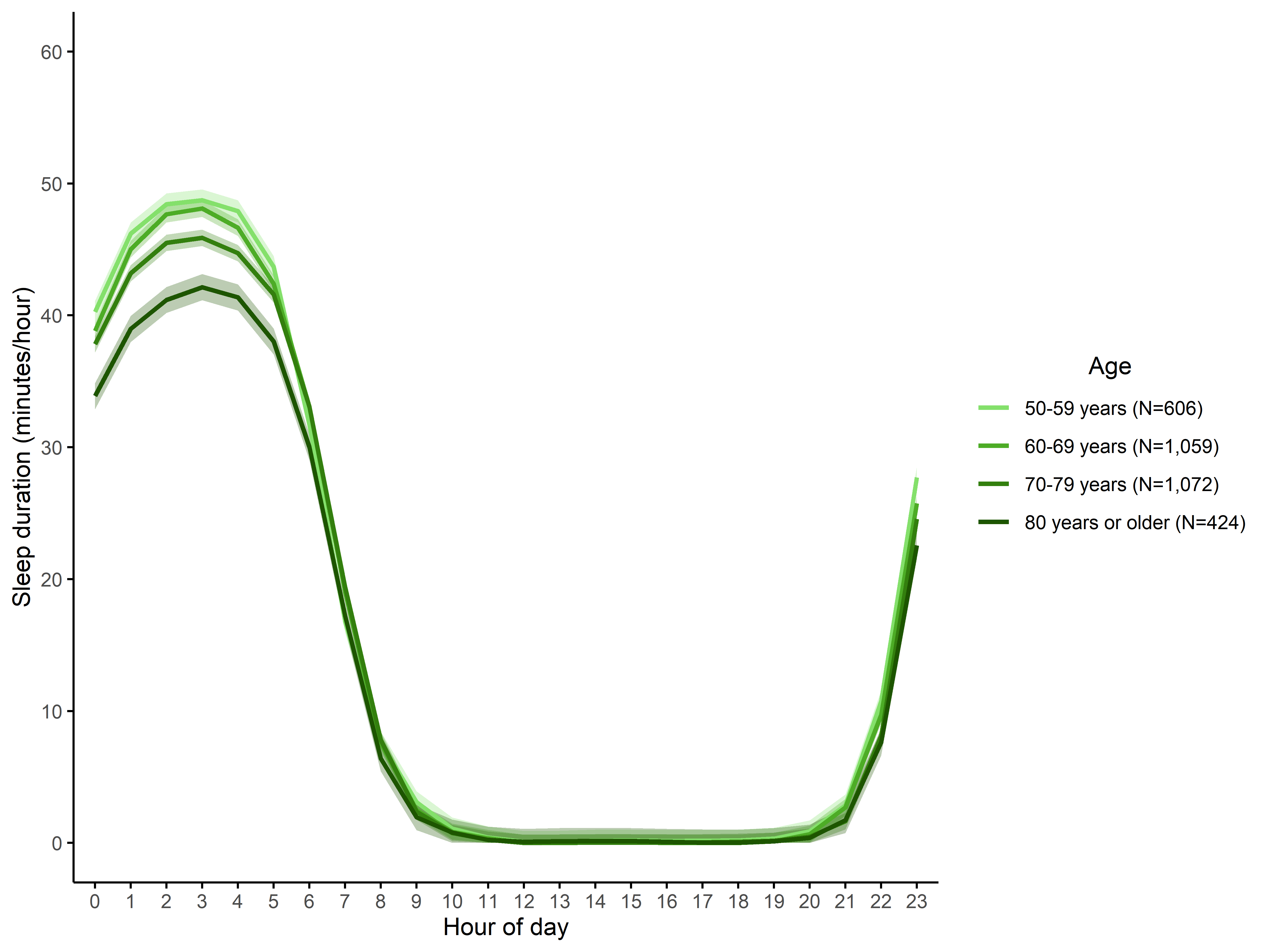
**Figure A5. 24-hour profile of sleep duration by age group (N = 3,161).**

Means were adjusted for sex and ribbon represents the 95% confidence interval.

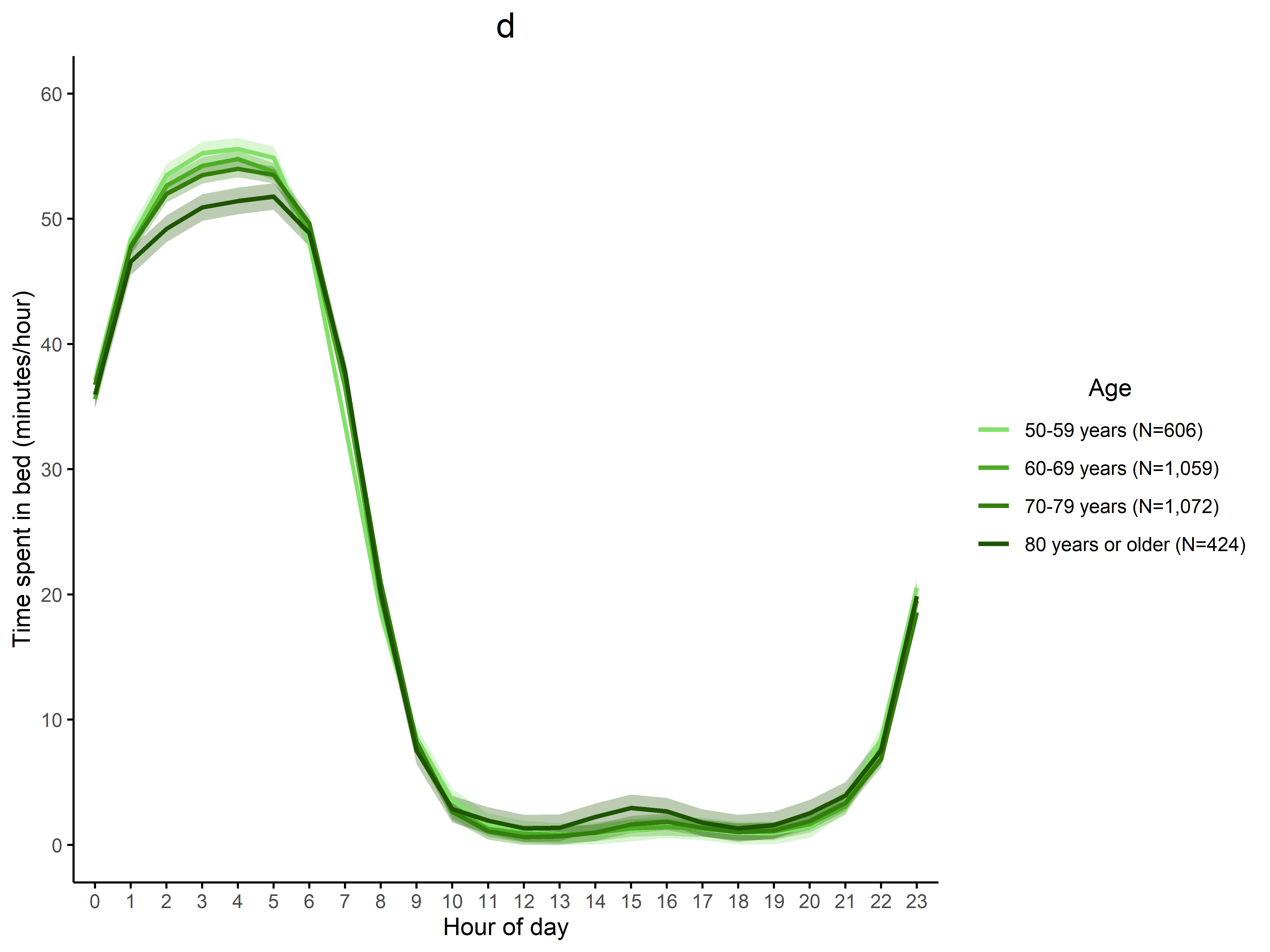

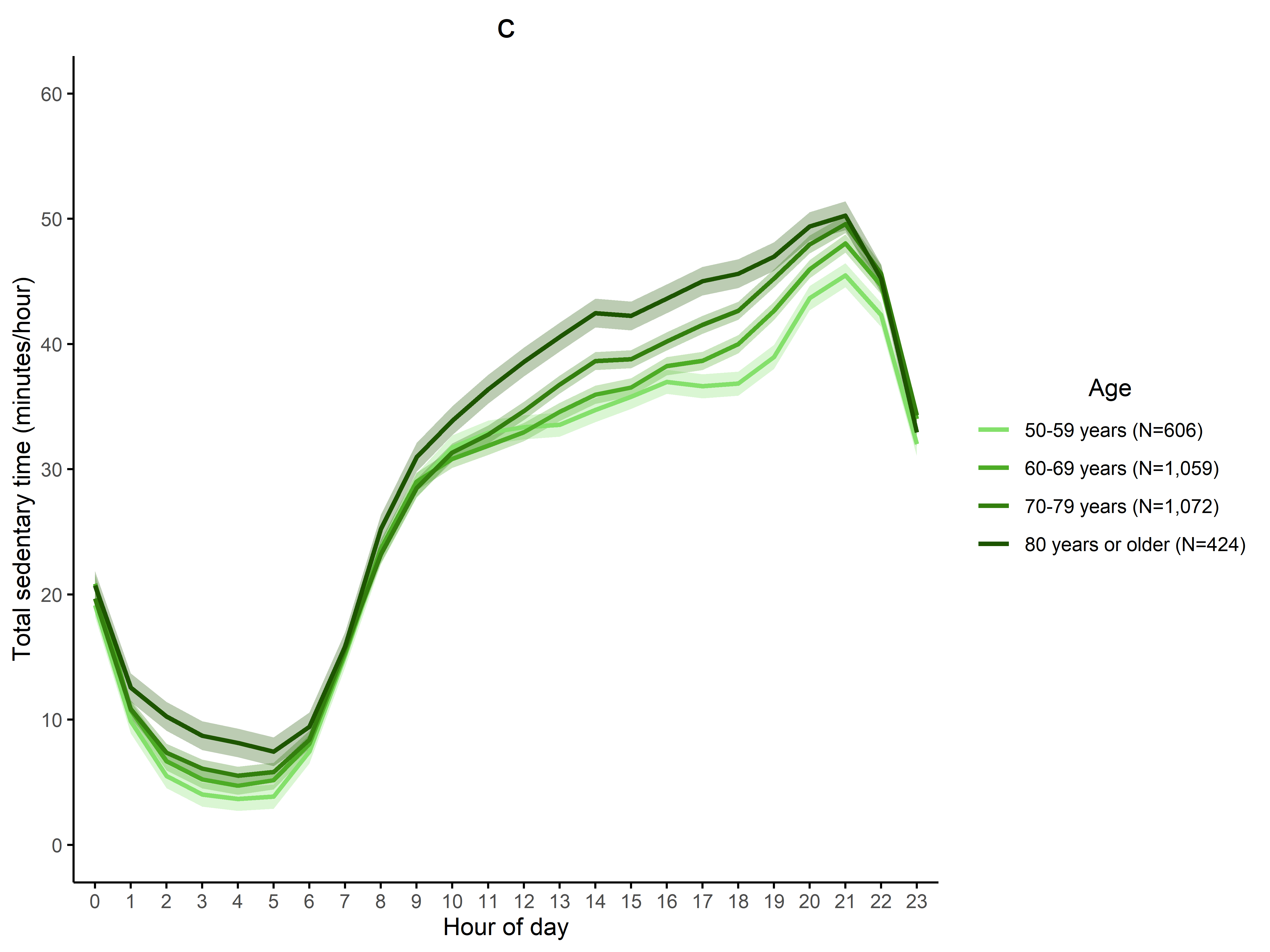

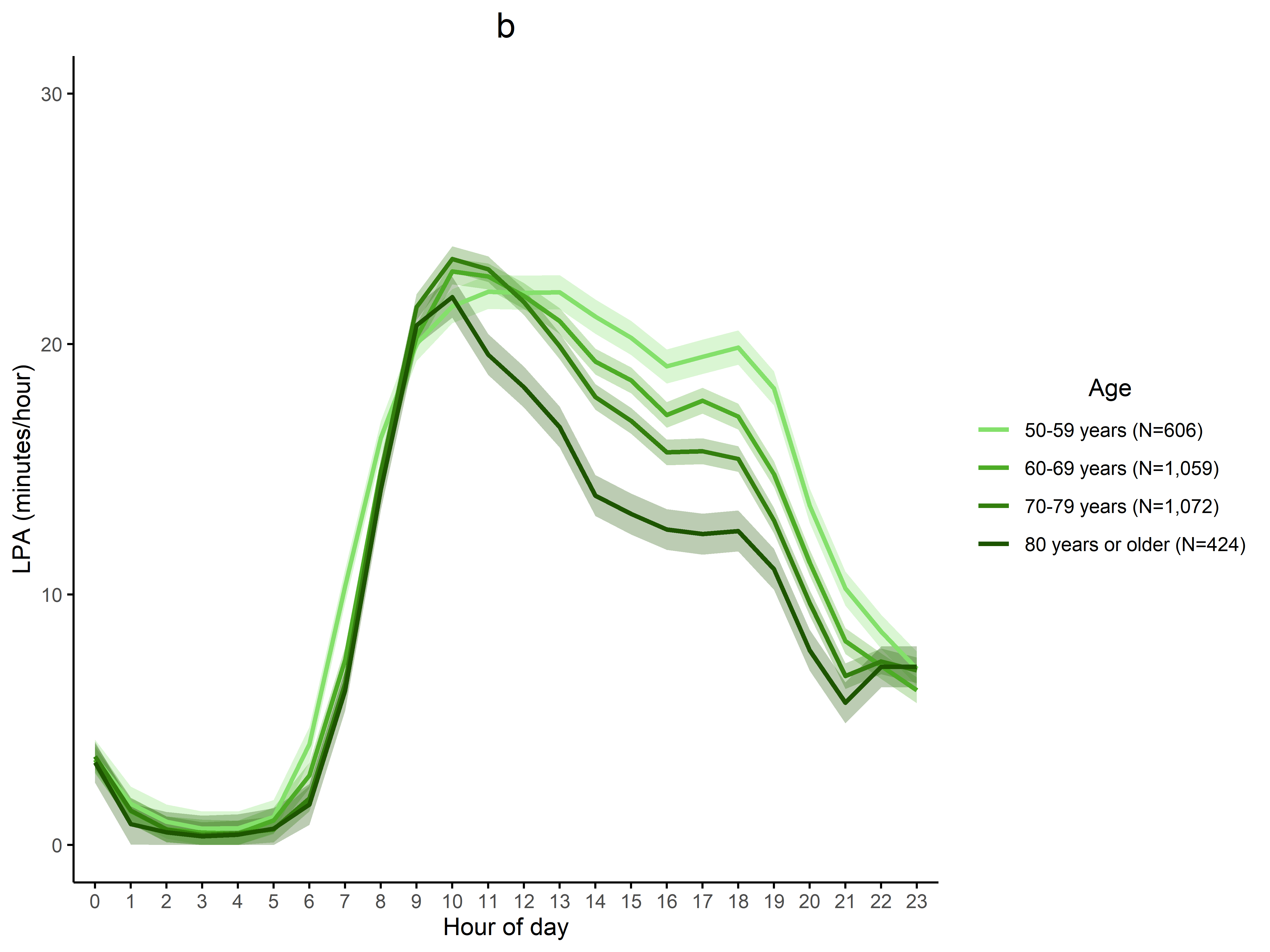

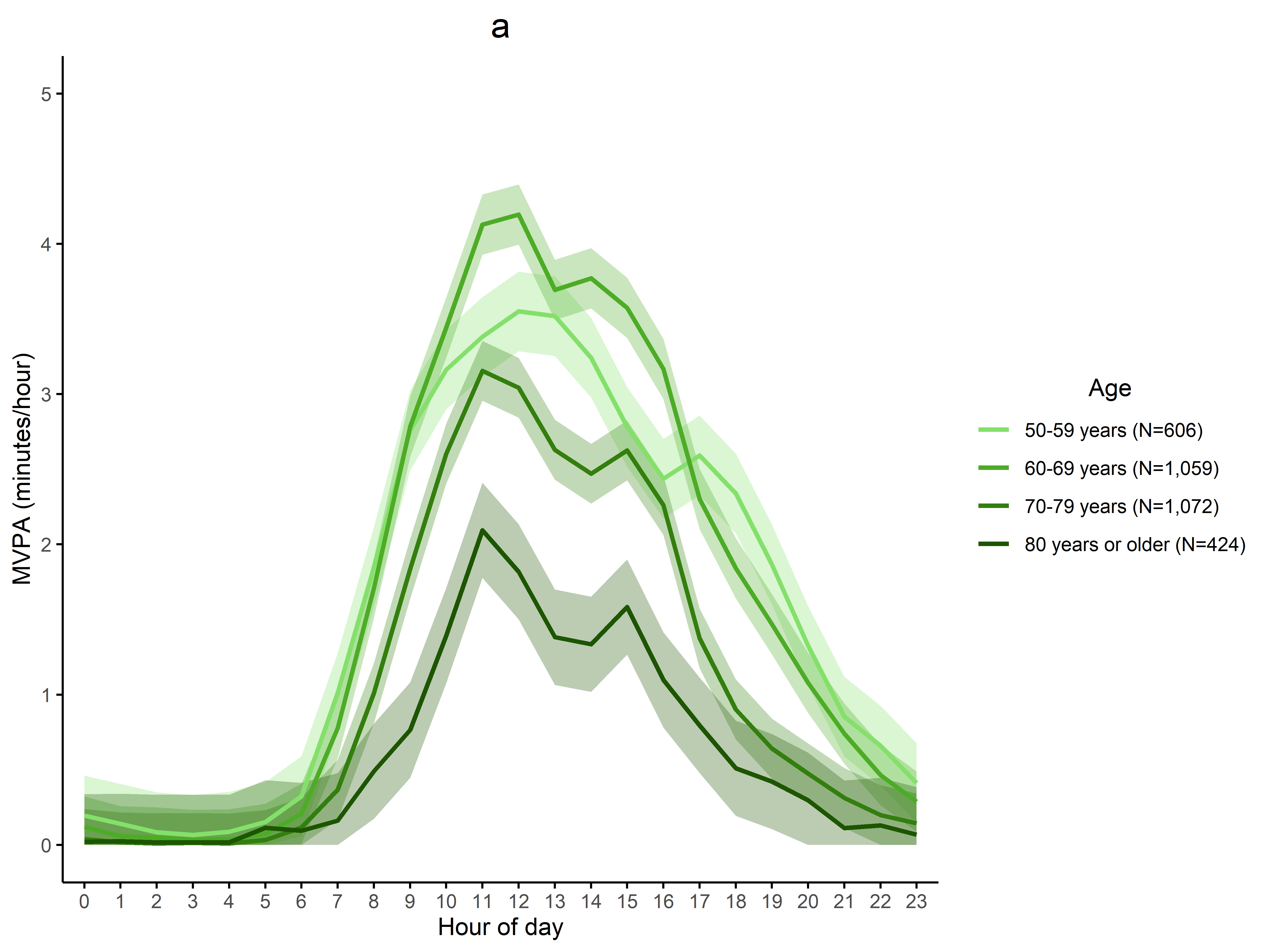

**Figure A6. 24-hour profile of MVPA (a), LPA (b), total sedentary time (c), and time spent in bed (d) by age group (N = 3,161).**

Means were adjusted for sex and ribbon represents the 95% confidence interval.

MVPA, moderate-vigorous physical activity; LPA, light physical activity.

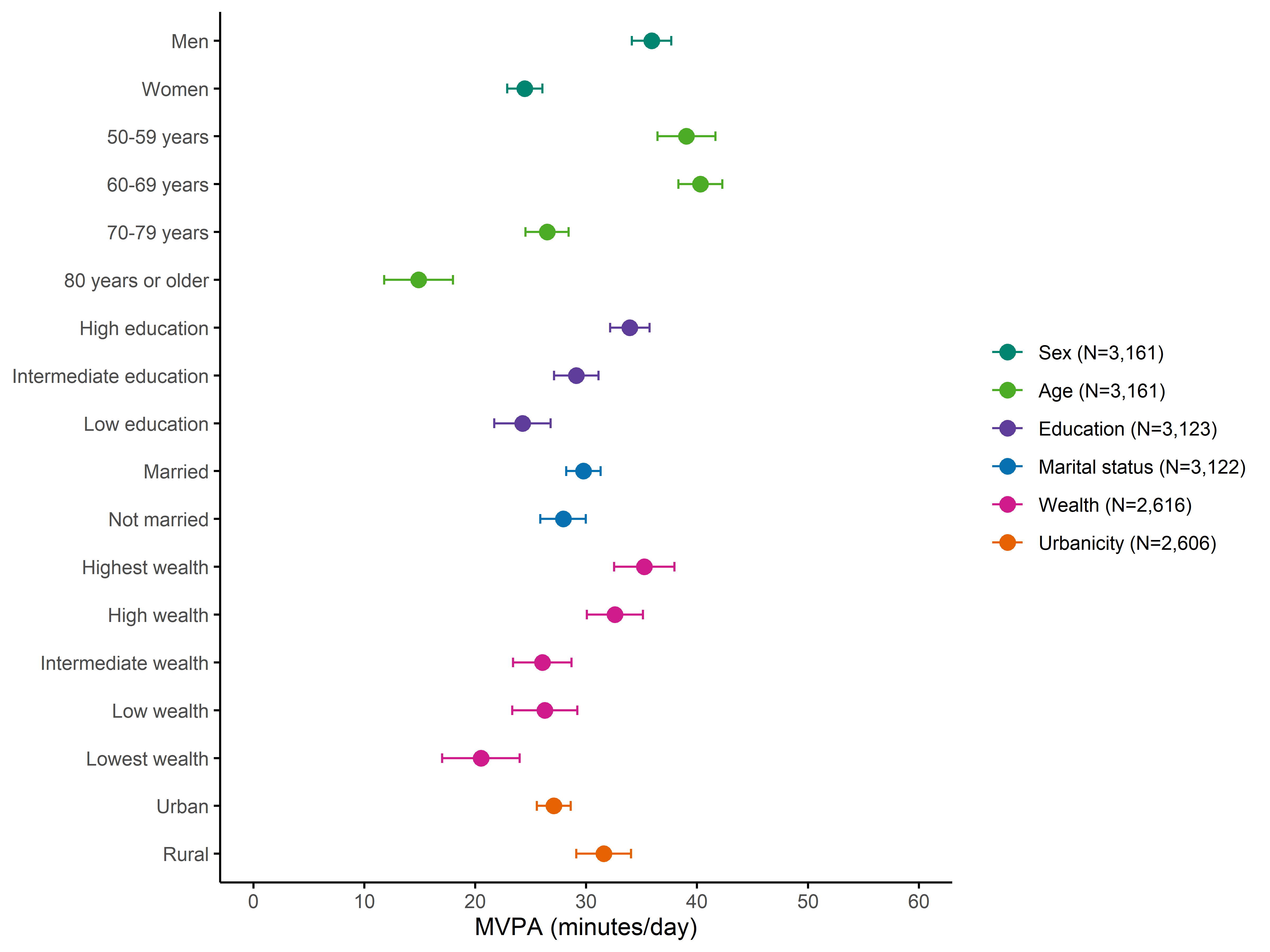
**Figure A7. Social, economic, and environmental disparities in time spent in moderate-to-vigorous physical activity (N = 3,161).**

The markers show marginal means and the horizontal lines show 95% confidence intervals.

Model 1: models for sex, age, and education were adjusted for sex and age group (where appropriate).

Model 2: models for marital status were additionally adjusted for education (Model 1 + education).

Model 3: models for wealth were additionally adjusted for marital status (Model 2 + marital status).

Model 4: models for urbanicity were additionally adjusted for wealth quintile (Model 3 + wealth).

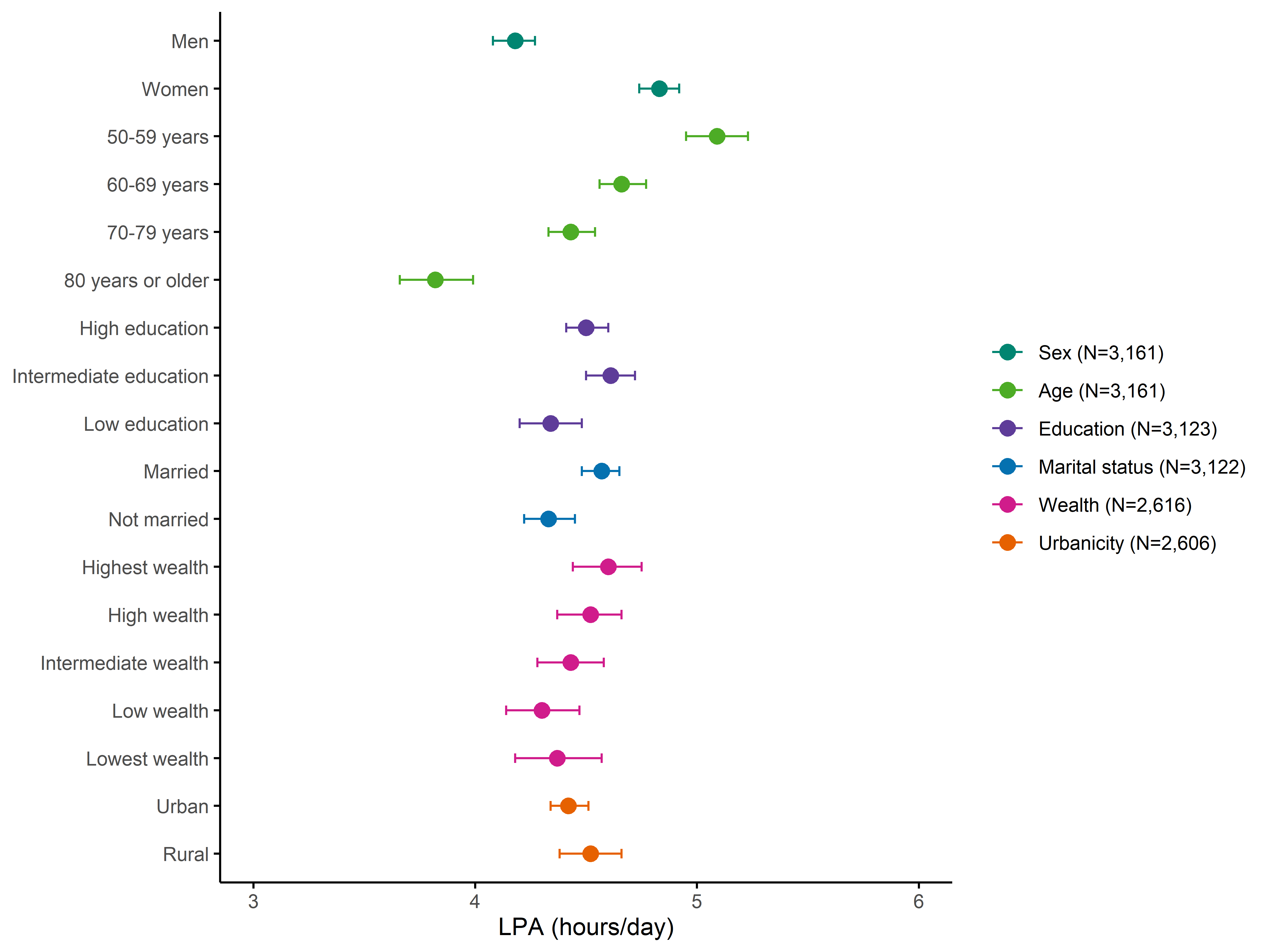
**Figure A8. Social, economic, and environmental disparities in time spent in light physical activity (N = 3,161).**

The markers show marginal means and the horizontal lines show 95% confidence intervals.

Model 1: models for sex, age, and education were adjusted for sex and age group (where appropriate).

Model 2: models for marital status were additionally adjusted for education (Model 1 + education).

Model 3: models for wealth were additionally adjusted for marital status (Model 2 + marital status).

Model 4: models for urbanicity were additionally adjusted for wealth quintile (Model 3 + wealth).

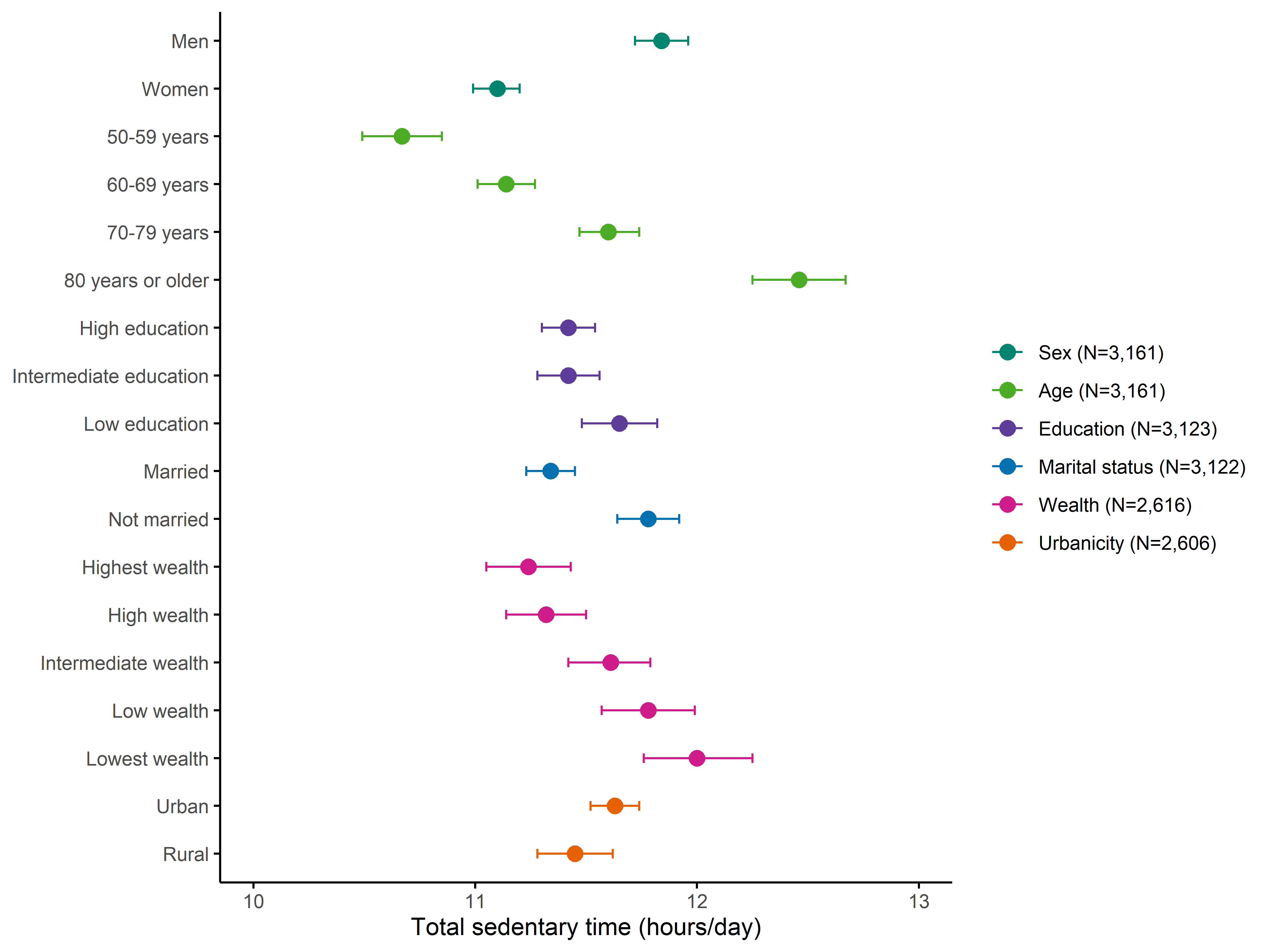
**Figure A9. Social, economic, and environmental disparities in total sedentary time (N = 3,161).**

The markers show marginal means and the horizontal lines show 95% confidence intervals.

Model 1: models for sex, age, and education were adjusted for sex and age group (where appropriate).

Model 2: models for marital status were additionally adjusted for education (Model 1 + education).

Model 3: models for wealth were additionally adjusted for marital status (Model 2 + marital status).

Model 4: models for urbanicity were additionally adjusted for wealth quintile (Model 3 + wealth).

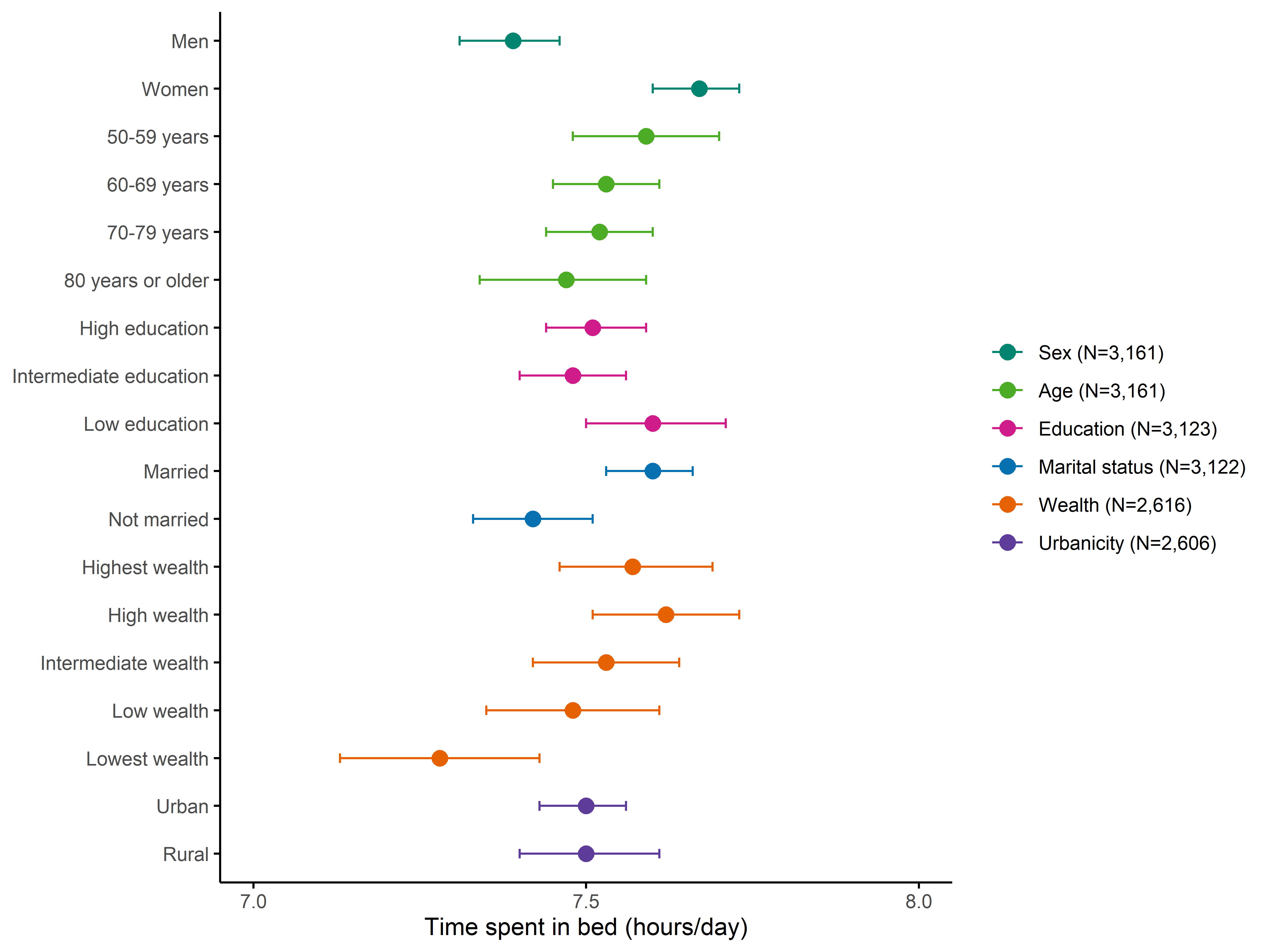
**Figure A10. Social, economic, and environmental disparities in time spent in bed (N = 3,161).**

The markers show marginal means and the horizontal lines show 95% confidence intervals.

Model 1: models for sex, age, and education were adjusted for sex and age group (where appropriate).

Model 2: models for marital status were additionally adjusted for education (Model 1 + education).

Model 3: models for wealth were additionally adjusted for marital status (Model 2 + marital status).

Model 4: models for urbanicity were additionally adjusted for wealth quintile (Model 3 + wealth).
